# HuBERT-ECG as a self-supervised foundation model for broad and scalable cardiac applications

**DOI:** 10.1101/2024.11.14.24317328

**Authors:** Edoardo Coppola, Mattia Savardi, Mauro Massussi, Marianna Adamo, Marco Metra, Alberto Signoroni

## Abstract

The electrocardiogram (ECG) is a widely accessible tool for cardiovascular assessment, and the growing availability of ECG datasets has enabled the emergence of ECG foundation models. However, such foundation models often lack extensive evaluation across clinically heterogeneous downstream tasks extending beyond conventional rhythm and conduction analysis. We present HuBERT-ECG, a self-supervised foundation ECG model pre-trained on 9.1 million 12-lead ECGs from four countries and diverse patient populations, and evaluated through fine-tuning on 21 independent datasets spanning more than 1.6k diagnostic and prognostic targets across adults and paediatric cohorts, including single-lead settings. These tasks cover conditions for which the ECG is the primary diagnostic modality, provides supportive but non-definitive diagnostic information, or enables acute-care prediction and prognostic modelling. Available in three model sizes to characterise scaling behaviour and support diverse computational constraints, HuBERT-ECG achieves AUROC ranging from 84% to 99% on ECG-primary diagnostic tasks, 76% to 97% on supportive diagnostic tasks, 74% to 91% on prognostic prediction tasks, and 88% to 92% on single-lead ECG benchmarks. Moreover, a large-scale multitask fine-tuning across 2.4 million subjects and 164 tasks simultaneously shows that AUROC further increases for clinically relevant tasks without extra task-specific supervision. We release pretrained models and code as building baselines.

## Introduction

The electrocardiogram (ECG) has long been a cornerstone of cardiovascular diagnostics, serving as a non-invasive, cost-effective and widely available tool for assessing cardiac and noncardiac diseases^1^. Through its recurring waveforms, the ECG encodes both physiological and pathological information through distinctive electrophysiological “fingerprints”^2^. Although structurally simple, these signals contain complex patterns that reveal deviations from normal cardiac function and provide valuable insights into cardiovascular health.

Integrating deep learning (DL) into ECG analysis represents a transformational opportunity to expand its clinical utility^3^. DL models can transform raw ECG traces into digital biomarkers for early detection, risk stratification, and intervention across a wide range of cardiovascular conditions. Previous studies have consistently shown strong performance in detecting conditions for which the ECG is the diagnostic gold standard (e.g. atrial fibrillation, tachycardia, and bradycardia)^4^. DL is also increasingly applied to diagnose morphological conditions primarily assessed through imaging, such as heart failure, pulmonary thromboembolism and aortic stenosis^5^. These studies suggest that ECG-based DL can capture signals associated with structural and systemic diseases beyond conventional electrophysiological abnormalities, including non-cardiac conditions such as liver disease and diabetes^6^, thereby complementing traditional imaging approaches^7^. Finally, predicting future cardiovascular events (CVEs) (e.g., myocardial infarction, stroke, death), represents one of the most challenging applications of ECG-based DL because of the difficulty of inferring long-term outcomes from ECG patterns alone^8–10^.

From a machine learning perspective, limited ECG dataset size, availability, annotation coverage and patient diversity have historically favoured highly specialised ECG models^11^ over versatile foundation models^12^. Such specialised models often struggle to generalise across domains or distributions^13^. For example, a model trained only to detect atrial fibrillation may fail to recognise other cardiac abnormalities, while one trained predominantly on older patients may not transfer well to younger populations. In stark contrast, foundation models pre-trained through self-supervision on large and diverse unlabelled datasets can learn robust and transferable representations adaptable to a wide range of downstream tasks with limited fine-tuning^14^.

Unimodal foundation models have transformed representation learning across natural language processing (e.g., the GPT series^15–18^, BERT^19^, T5^20^), computer vision (e.g., DINO^21^, MAE^22^), and audio processing (e.g., Wav2Vec2^23^, HuBERT^24^) through large-scale self-supervised pre-training on unannotated data. More recently, these advances have enabled multimodal systems integrating heterogeneous data modalities. In medical domains, progress has been slower because of limited large-scale datasets^14,25^, but foundation models have begun emerging across computational pathology (e.g., CONCH^26^, Prov-GigaPath^27^, Virchow^28^), radiology (KAD^29^, CheXzero^30^), medical image segmentation (MedSAM^31^), echocardiography (EchoCLIP^32^), and broader biomedical tasks BiomedGPT^33^. Electrocardiography has also witnessed the emergence of foundation-model-oriented approaches. However, current ECG foundation models often lack one or more key ingredients for truly foundational design, including geographically diverse and large-scale pre-training data, self-supervised optimisation, and, most importantly, extensive evaluation across multiple datasets and clinically heterogeneous downstream tasks, particularly in low-data and non-standard settings. Several works^34–39^ leverage contrastive-learning, but lack the combination of large-scale, diverse pre-training and extensive evaluation. HeartBEiT^40^ exploits self-supervision during large-scale pre-training but suffers from limited source diversity and narrowly scoped evaluation (only three tasks). KED^41^ demonstrates impressive zero-shot capabilities via text supervision, yet lacks large-scale, diverse pre-training, omits self-supervision (reaching less general representations^21^) and is evaluated only on form and rhythm conditions. Most recently, ECGFounder^42^ offers detection capabilities for 150 conditions through supervised training on 10 million ECGs from a single site.

In this work, inspired by HuBERT’s architecture^24^, we present HuBERT-ECG, a self-supervised ECG foundation model designed for broad transferability across a wide range of clinically heterogeneous tasks, from ECG interpretation to cardiovascular risk prediction. HuBERT-ECG is pre-trained on an extensive dataset of 9.1 million 12-lead ECGs from four countries and diverse patient populations. Nearly 3 million ECGs in our dataset are annotated for one or more conditions from a comprehensive set of more than 1.6k diagnostic and prognostic target spanning electrophysiological findings, structural and functional cardiac abnormalities, and clinical outcomes. These annotations originate from publicly available standard ECG benchmarks and encompass both ECG-defined findings and clinically established diagnostic or prognostic labels associated with the recorded ECGs. To systematically characterise downstream transferability, we group evaluation tasks into three categories: type-1, where ECGs are the primary diagnostic modality (e.g., atrial fibrillation, bundle branch blocks, ST-segment abnormalities); type-2, where ECGs provide supportive but non-definitive diagnostic information (e.g., heart failure, ventricular hypertrophy, myocardial infarction, valvular disease); and type-3, where ECGs provide signals of future CVEs or clinical outcomes (e.g., mortality, hospitalisation, acute-care deterioration).

Through supervised finetuning, HuBERT-ECG is evaluated on 21 independent datasets of various size and population diversity spanning a wide range of tasks including ECG-defined finding detection, structural and functional cardiovascular assessment, clinical outcomes, acute-care prediction, short- and long-term prognostic modelling, paediatric ECG interpretation, and portable device-derived single-lead ECG analysis. Beyond independent dataset evaluations, to assess whether joint finetuning on multiple tasks can increase performance observed for task-specific training, we further explore large-scale multitask finetuning through a harmonised training framework integrating 164 overlapping cardiovascular labels and 2.4 million subjects across public ECG datasets. This overall comprehensive evaluation is conducted for three model sizes—SMALL (30M), BASE (97M) and LARGE (188M)—to investigate performance scaling behaviour and support diverse computational constraints. Finally, we compare retrospectively HuBERT-ECG’s with human annotators and analyse the efficiency of our fine-tuning protocol to guide future users.

Across these diverse evaluation scenarios, HuBERT-ECG demonstrates strong and consistent performance on both ECG-primary diagnostic tasks and more challenging supportive diagnostic and prognostic settings, while also showing robust transferability across datasets, populations, and acquisition modalities. Beyond this, our large-scale multitask finetuning further demonstrates that jointly learning heterogeneous cardiovascular tasks can improve performance even for specific target conditions without requiring additional task-specific supervision—a finding particularly relevant for clinically difficult tasks where labelled data remain scarce or expensive to curate. These findings, together with performance superior to that of human annotators and proven fine-tuning efficiency, position HuBERT-ECG as a broadly transferable foundation model for cardiovascular representation learning. To support future research, we publicly release pretrained models and code.

## Results

### ECG Representation Learning through Self-supervised Pre-training

To develop a robust ECG foundation model, we propose HuBERT-ECG, a self-supervised foundation model learning meaningful representations through an iterative process combining (1) feature extraction, (2) clustering and (3) masked modelling. Specifically, descriptive ECG features are extracted from ECG segments and then clustered by a clustering model. Subsequently, during pre-training, embedded versions of ECG segments are masked and HuBERT-ECG is trained to predict which cluster they belong to. By repeating this process across multiple pre-training iterations, and progressively refining the feature extraction, HuBERT-ECG improves at capturing the underlying structure of ECG signals, ultimately achieving generalisable ECG representations.

We pre-train HuBERT-ECG on an extensive and diverse dataset of 9.1 million ECGs from four countries (Fig. 1a-d, Fig. 2a, “Methods” – *Data and Preprocessing*). In the first iteration, before masked modelling, we extract and cluster Mel Frequency Cepstral Coefficients^43^ (MFCCs) from raw ECG segments. MFCCs, capture key frequency components and highlight essential ECG structure. In the second iteration, we extract semantically higher features from the model’s 8th layer (latent representations) and refine clusters. This iterative refinement allows HuBERT-ECG to learn a deeper understanding of ECG patterns and structure through masked modelling. HuBERT-ECG architecture and pre-training specifics are detailed in “Methods” - *HuBERT-ECG Architecture and Theoretical Framework*. Feature extraction and clustering are illustrated in “Methods” – *Unsupervised pseudo-label discovery*. Specifications and implementation details are provided in “Methods” – *Implementations* and “Methods” – *Self-supervised pre-training*.

**Fig 1.**
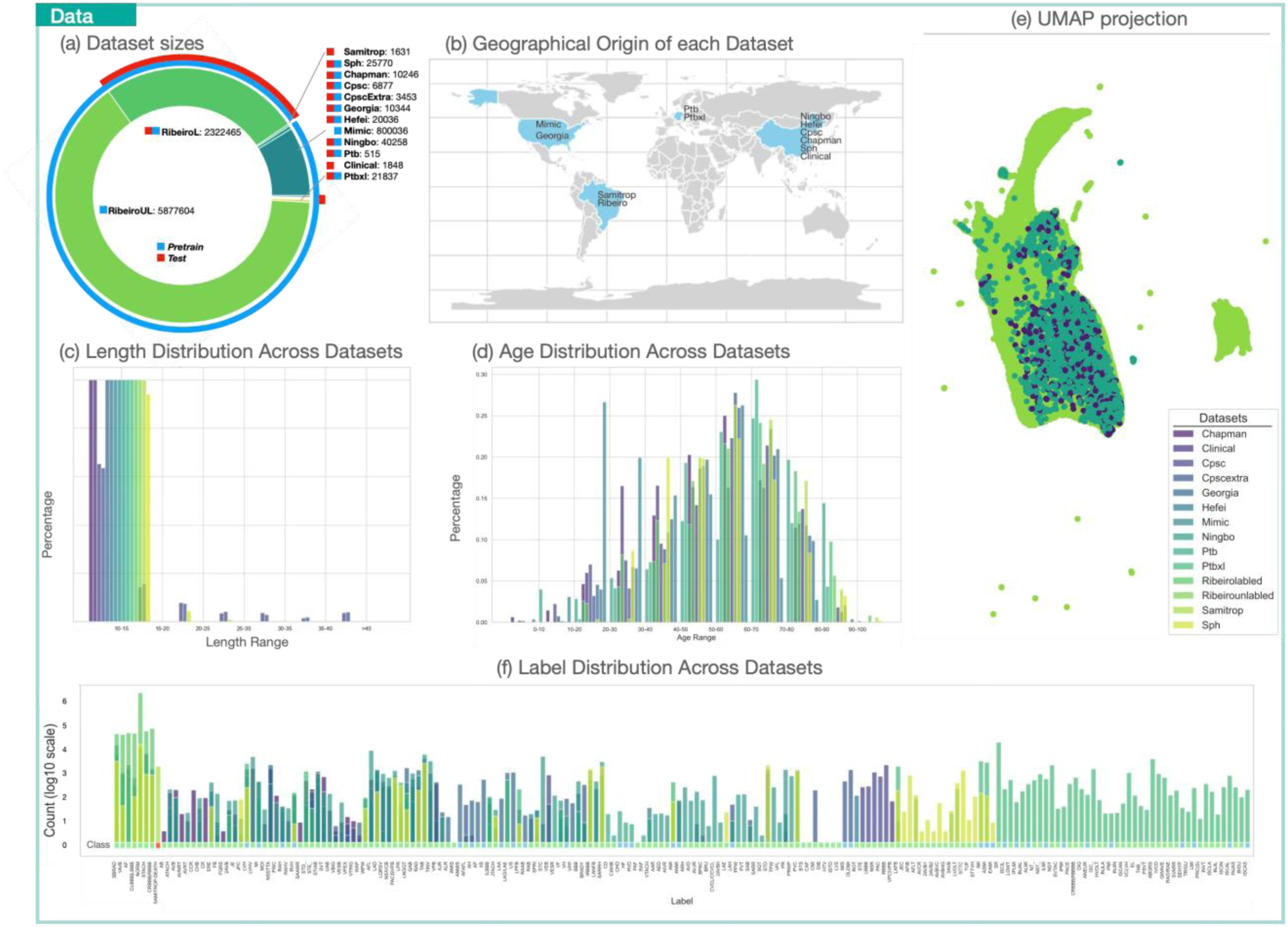
General information on the 9.1 million ECG dataset. **(a)** Used dataset and their sizes. The blue bar highlights the dataset used for the pre-training while the red one is the testing one. **(b)** Geographical origin of the considered datasets. **(c)** ECG length in seconds. **(d)** Age distribution. **(e)** UMAP projection (uniform sample of 30% of majority of datasets for the sake of visualisation). **(f)** Label distribution and grouping according to 3 classes (see the coloured bar below the distribution): Task group 1: green colour for tasks in which the ECG the primary diagnosis tool; task group 2: blue colour for tasks in which the ECG is not the primary diagnostic tool but a potentially informative support; task group 3: red colour for tasks for which the ECG is used to predict future CVEs. EchoNext, MITHSDB, ZZU-pECG, and MIMIC-IV-ECG are not reported in this figure to avoid visual clutter but have been described in Table 7 and Supplementary Tables 2-3. Label abbreviations and corresponding diagnosis are reported in Supplementary Table 1.

**Fig 2.**
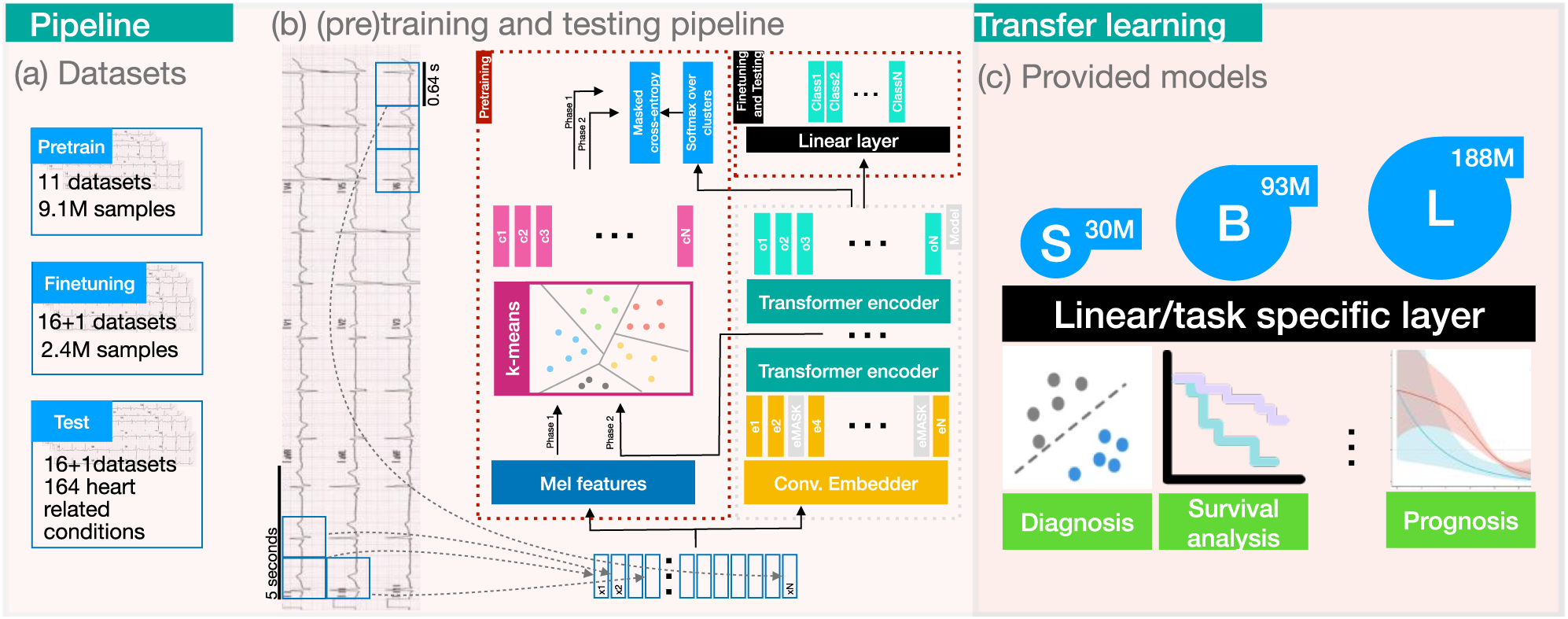
An overview of the proposed deep learning pipeline. (a) **Datasets**: The model is pre-trained on 11 datasets comprising over 9.1 million samples. Then, the model is fine-tuned and tested on 16 datasets and their aggregation covering 164 heart-related conditions. (b) **Pre-training and Testing Pipeline**: The pipeline begins with Mel feature extraction, followed by k-means clustering, and includes two training phases with masked cross-entropy loss and softmax classification. A convolutional embedder and a Transformer encoder process the data, culminating in a linear output layer for classification. (c) **Transfer Learning and Provided Models**: Three model variants (**S**MALL, **B**ASE, **L**ARGE) with increasing parameter counts (30M, 93M, and 188M) are pre-trained, tested and, eventually, provided to enable transfer learning for many downstream tasks such as diagnosis, survival analysis, and prognosis. Fine-tuning can be performed by simply adding a task-specific linear layer.

To investigate performance scaling behaviour and support diverse computational needs, we also pre-train SMALL and LARGE configurations in a single iteration using more refined latent representations.

### Supervised fine-tuning on downstream data and evaluation

To comprehensively assess HuBERT-ECG’s transferability across clinically heterogeneous tasks, we compile 21 downstream datasets for fine-tuning and evaluation. These datasets include: the labelled partition of Ribeiro^44^ (also known as CODE); CPSC and CPSC-Extra^45^; PTB^46^; PTB-XL^47^ which, with 6 different sets of conditions, gives rise to 6 different datasets; the publicly available partition of Georgia^48^; Chapman-Shaoxing^49^ (Chapman); Ningbo First Hospital^50^ (Ningbo); the public partition of the Tianchi Arrhythmia Competition^a^ dataset (Hefei); Shandong Provincial Hospital^51^ (SPH); SaMi-Trop^52^; the so called “Clinical data” from the work of Tian et al.^41^; EchoNext^53^, ZZU-pECG^54^; MIMIC-IV-ECG^55^; and MIT Heart Sound DB (MITHSDB) ^56^. In addition, to investigate gains from joint optimisation for numerous tasks over task-specific training, we harmonise labels from each dataset (except “Clinical data”, EchoNext, ZZU-pECG, MIMIC-IV-ECG and MITHSDB; see “Methods” – *Data and Preprocessing* for motivations about this exclusion) and merge all sources into a comprehensive dataset named *Cardio-Learning*. This dataset includes 2.4 million ECGs from four countries and diverse populations, each labelled with one or more conditions from a highly imbalanced set of 164 targets. This data source covers all task types. For terminological clarity, in this work we use the term *task* to denote a specific learning target (e.g., detecting atrial fibrillation, identifying left ventricular hypertrophy, or predicting mortality risk). For single-label datasets (SaMi-Trop, MITHSDB), each dataset corresponds to one task. For multi-label datasets (all the other datasets), each label is predicted independently in a one-vs-rest setting, so a dataset with L labels comprises L distinct tasks, each shaped by its own prevalence, population, and noise.

After fine-tuning, HuBERT-ECG is evaluated on hold-out test sets of 22 datasets (21 individual + Cardio-Learning). For 17 highly diverse datasets, fine-tuning performance is compared with that achieved by training the same model from scratch, i.e., using randomly initialised weights. This comparison reveals the net contribution of pre-training over training from scratch, showing that the fine-tuned HuBERT-ECG consistently outperforms its randomly initialised counterpart in efficiency and generalisation. Given this consistency, we report results on the remaining minority of datasets only after finetuning to prioritise breadth of clinical assessment.

To ensure consistent evaluation, address label distribution disparities, and employ a robust, threshold-independent metric, we report model performance using the Area Under the Receiver Operating Characteristic Curve (AUROC) for classification tasks, pointing out that different AUROC ceilings may be expected for different tasks. For regression tasks we use z-normalized Mean Absolute Error (MAE). (see “Methods” – *Evaluation Metrics*). The fine-tuning procedure is detailed in “Methods” - *Supervised Fine-tuning*. The following results, presented across diverse application scenarios, are meant to demonstrate the broad robustness and transferability of HuBERT-ECG’s self-supervised ECG representations.

#### HuBERT-ECG is efficient on small-sampled datasets

ECG-based interpretation is affects by limited dataset size, and PTB (515 instances, 17 tasks), CPSC (6878 ECGs, 9 tasks), CPSC-Extra (3453 ECGs, 52 tasks), and “Clinical data” (1784 ECGs, 12 tasks) are clear examples. Here, HuBERT-ECG consistently shows solid performance and exhibits distinct macro-trends. On PTB (Fig. 3a), the SMALL and BASE configurations perform comparably well when fine-tuned, achieving AUROCs of 0.843±0.029 and 0.848±0.03, respectively. On CPSC (Fig. 3b), the SMALL model size achieves an AUROC of 0.945±0.002, slightly less than BASE (0.949±0.002) and LARGE (0.949±0.006) models. On this dataset we outperform contrastive learning approaches and foundation models (e.g., Liu et al.^36,37^, Tian et al.^41^ and Zhang et al.^39^ achieved AUROCs of 0.932, 0.915, 0.910, and 0.920, respectively) while remaining competitive with models specifically pre-trained and optimised for arrhythmia detection only (e.g., Na et al.^57^ with 0.980, Zhou et al.^58^ with 0.965). On CPSC-Extra (Fig. 3c), a more challenging dataset, HuBERT-ECG SMALL performs best (0.894±0.076), even surpassing ECGFounder when fine-tuned on this source (AUROC=0.852). Conversely, larger configurations perform less effectively (BASE 0.756±0.02; LARGE 0.774±0.023). This suggests the advantages of smaller architectures (HuBERT-ECG SMALL and ECGFounder parameter count ∼ 30M) in multi-label, low-data contexts. On “Clinical data” (Fig. 3d), the BASE configuration stands at top (0.915±0.009), followed by LARGE (0.910±0.015) and SMALL (0.893±0.017). Across these datasets, fine-tuning improves upon training from scratch by 3–8% AUROC points.

**Fig. 3.**
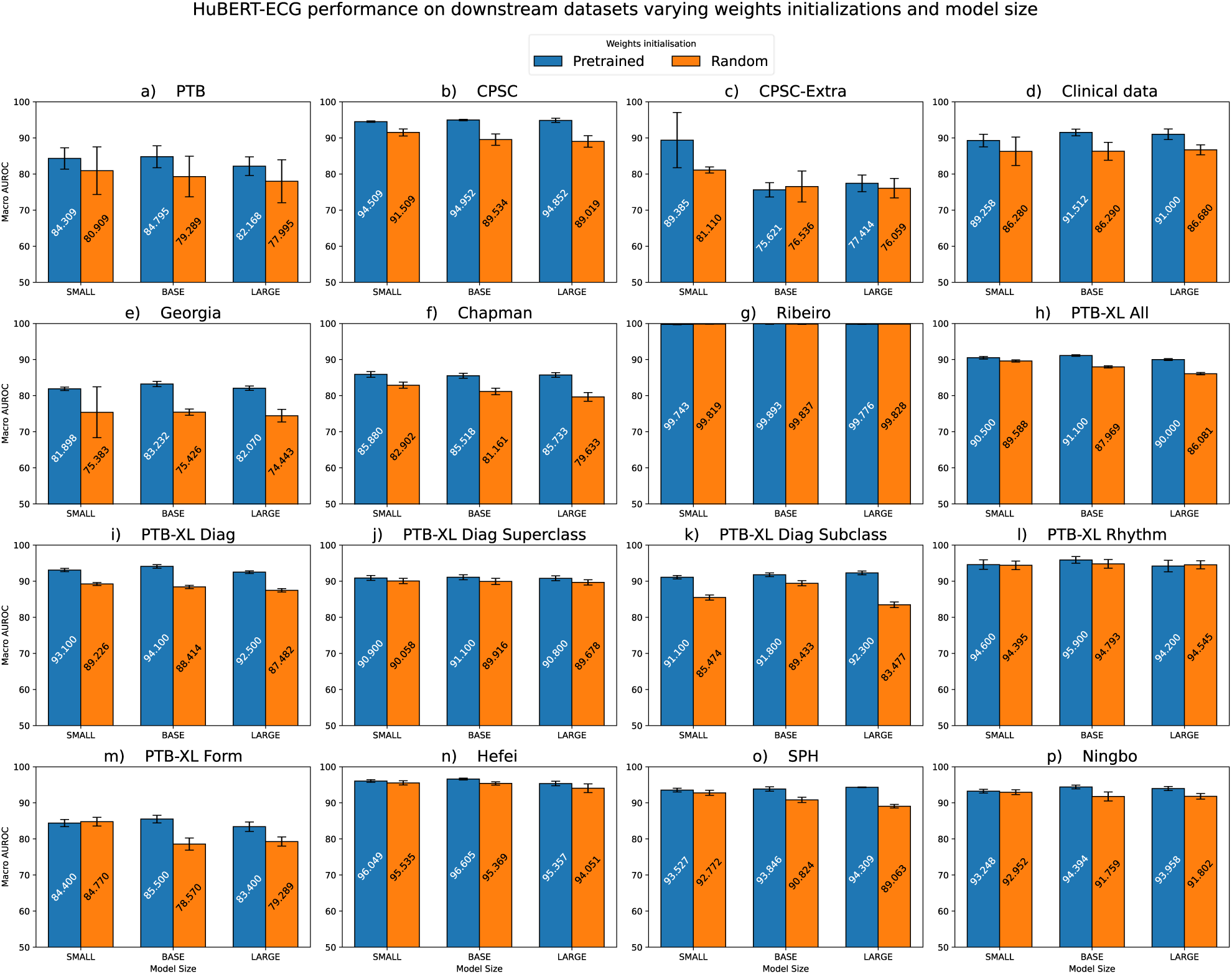
HuBERT-ECG macro-averaged performance across 16 downstream datasets varying weights initialization and model size. Performance uncertainty on datasets with no predefined test set is reported through standard deviations across fold performance, while on datasets with predefined test sets uncertainty is assessed using 95% confidence intervals obtained via empirical bootstrapping with 1000 bootstrap iterations.

#### HuBERT-ECG remains efficient in diagnostically difficult contexts

ECG-based diagnostic complexity is also influenced by large label spaces, low per-class prevalence and many overlapping diagnostic statements. Georgia (10k ECGs, 65 tasks) and Chapman (10k ECGs, 52 tasks) exemplify these factors. On Georgia (Fig. 3e), HuBERT-ECG BASE tops alternatives (AUROC=0.832±0.007). For the sake of comparison, Tian et al.^41^ and Zhang et al.^39^ facilitated the learning problem by limiting the diagnostic scope to 20 and 46 classes, reaching AUROCs of 0.900 and 0.866. When we fine-tune HuBERT-ECG BASE on these subsets, we substantially match competitors: 0.895±0.079 on the 20-label problem, 0.858±0.093 on the 46-label one. Conversely, when directly evaluated on these condition subsets, despite being fine-tuned to detect all 65 conditions simultaneously, HuBERT-ECG BASE remains competitive (AUROC scores: 0.853±0.003, 0.855±0.093). On Chapman (Fig. 3f), HuBERT-ECG SMALL slightly tops larger configurations (0.859±0.008). To address label distribution imbalances, Chapman’s authors suggested grouping conditions into four macro-categories, highly simplifying the diagnostic task. Following this, Liu et al.^36^ and Kiyasseh et al.^34^ reported AUROCs of 0.997 and 0.932, respectively. When HuBERT-ECG BASE is fine-tuned under the same grouping, it matches the state-of-the-art on this dataset (0.995±0.001). Notably, testing directly all configurations on these four broader classes—despite being fine-tuned to recognise the original 52—delivers highly competitive AUROC scores (SMALL 0.975±0.002; BASE 0.975±0.009; LARGE 0.984±0.001) without marked performance loss. This strong performance underscores HUBERT-ECG’s robustness across varying levels of diagnostic granularity, even without simplifications of the underlying learning problem.

#### HuBERT-ECG enables accurate large-scale ECG-based diagnostics

We next evaluate HuBERT-ECG on Ribeiro, PTB-XL, Hefei, SPH and Ningbo, large-scale datasets involving tens of thousands to millions of patients and broad diagnostic spaces.

On Ribeiro (2.3 million ECGs, 6 tasks), all HuBERT-ECG configurations excel regardless of weight initialization (Fig. 3g), with fine-tuned BASE performing best (AUROC=0.9989). Table 2 (BASE only for conciseness), shows consistent gains over Ribeiro et al.^44^ and ECGFounder^42^ across sensitivity, specificity, AUROC, and AUPRC.

**Table 1.** HuBERT-ECG architecture summary.

|  |  | SMALL | BASE | LARGE |
| --- | --- | --- | --- | --- |
| Convolutional embedder | strides | 4, 2, 2, 2, 2 |  |  |
|  | kernels width | 10, 3, 3, 2, 2 |  |  |
|  | channels | 512 |  |  |
| Transformer encoder | layers | 8 | 12 | 16 |
|  | internal dimension | 512 | 768 | 960 |
|  | feed-forward dimension | 2048 | 3072 | 3840 |
|  | attention heads | 8 | 8 | 12 |
| Embedding dimension W |  | 256 | 256 | 512 |
| Number of parameters |  | 30M | 93M | 188M |

**Table 2.**
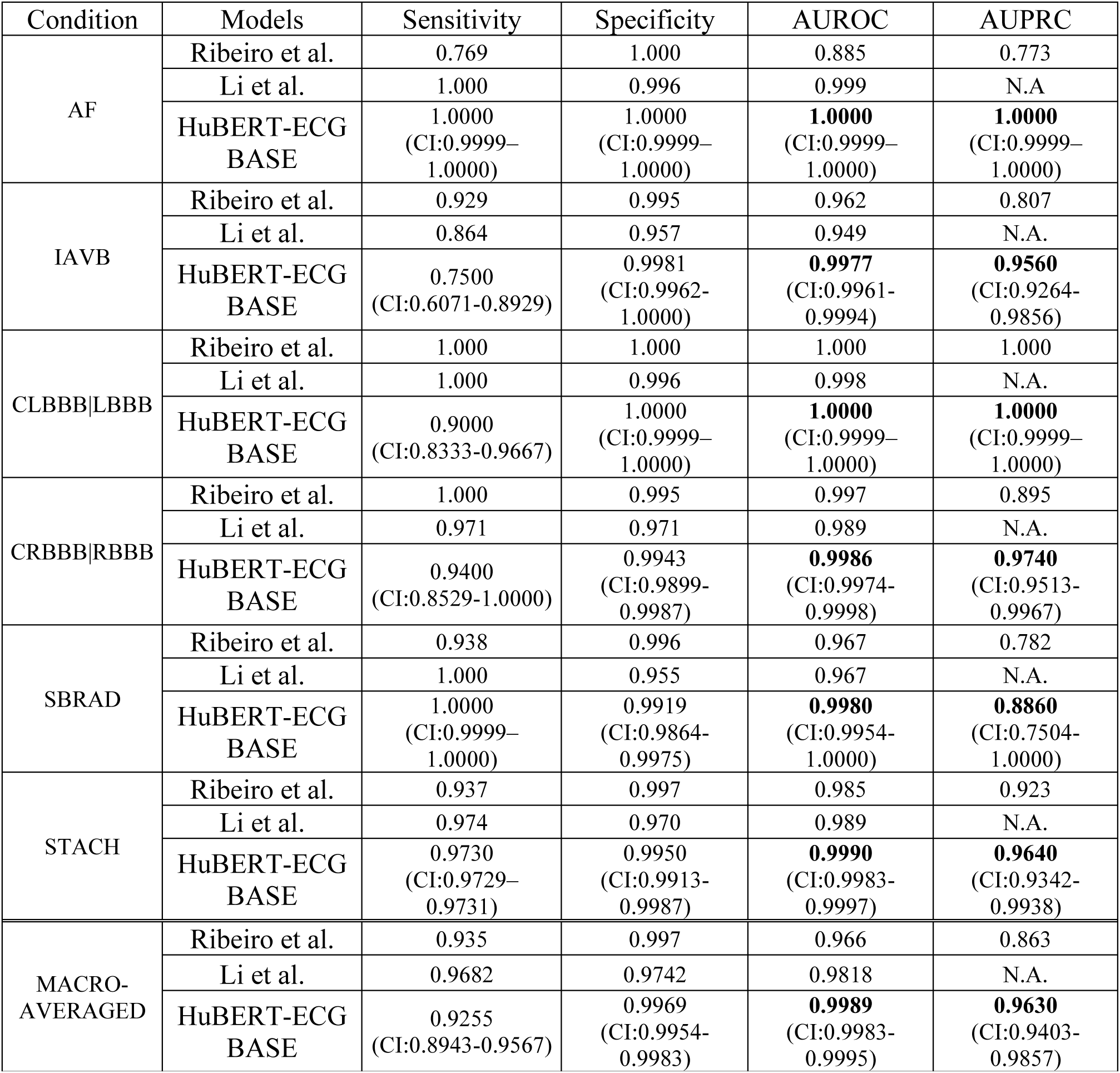
Fine-tuned HuBERT-ECG BASE performance on Ribeiro benchmarked against that from Ribeiro et al. ^44^ **and Li et al.**^42^ **according to multiple metrics.** Sensitivity and Specificity are obtained setting the threshold to 0.5, as done by Ribeiro et al., for a fair comparison. Performance uncertainty is reported using 95% confidence intervals obtained via empirical bootstrapping using 1000 bootstrap iterations. Label abbreviations and corresponding diagnosis can be found in Supplementary Table 1. A full comparison of all HuBERT-ECG model sizes, which may perform better than the BASE one on certain conditions for certain metrics, is provided in Supplementary Table 7.

PTB-XL^47^ (21837 ECGs, 71 tasks) cover diagnostic, form and rhythm annotations that can be grouped into 44 diagnostic conditions, 23 diagnostic subclasses, 5 broader diagnostic superclasses, 12 rhythm statements, and 19 form abnormalities. This gives rise to 6 different datasets, known as PTB-XL *All*, *Form*, *Rhythm*, *Diagnostic*, *Diag. Subclass*, *Diag. Superclass*. On these well-known benchmarks (Fig. 3h-m), HuBERT-ECG BASE exceeds 0.900 AUROC on 5 out of 6 datasets. More generally, HuBERT-ECG’s dataset-agnostic pretraining enables competitive performance with state-of-the-art works tailored on PTB-XL^59^, and improvements upon other solutions^36,37,39^ (Supplementary Table 9 for extensive comparisons). Compared to foundation models like KED^41^, ECGFounder^42^ and ECGFM^39^—which restricted their evaluation scope (just 46 diagnostic classes for KED and 30 for the others) and reported AUROCs of 0.858, 0.924 and 0.917, respectively—HuBERT-ECG shows markedly superior performance, even under full 71-label fine-tuning. Under matched setups, HuBERT-ECG BASE reaches superior AUROCs: 0.911±0.003 (46-label problem), 0.921±0.009 (30-label problem). Even when fine-tuned on all 71 labels, it still achieves strong transfer (46-label AUROCs: 0.900 SMALL, 0.904 BASE, 0.901 LARGE; 30-label AUROCs: 0.918 SMALL, 0.926 BASE, 0.923 LARGE), indicating better generalisation than prior foundation models.

On Hefei (20k+ ECGs, 29 conditions), BASE performs best (AUROC=0.9661±0.0025, Fig. 3n), outperforming fine-tuned ECGFounder (0.9532) and KED (0.9490).

On SPH^51^ (25,770 ECGs, 44 labels), all variants perform well, with LARGE at top (AUROC=0.943, Fig. 3o). Clinically relevant prior work^60^ concentrated only on detecting four myocardial infarction subtasks—AMI, IMI, ASMI, EAMI (see Supplementary Table 1 for abbreviations)—reaching AUROCs of 94.68%, 92.73%, 92.55%, 99.86%, respectively. Even under SPH’s full complexity, fine-tuned HuBERT-ECG BASE reaches 95.79%, 93.94%, 94.58%, 99.83%, with SMALL performing marginally better (AMI=94.25%, IMI=93.87%, ASMI=96.81%, EAMI=99.81%), and LARGE best (AMI=96.83%, IMI=95.13%, ASMI=97.48%, EAMI=99.96%).

On Ningbo^50^ (34,905 ECGs, 76 labels), fine-tuned BASE performs best (AUROC=0.9439±0.0053, Fig. 3p), surpassing KED (0.9382) and competing with ECGFounder (0.9531).

##### HuBERT-ECG transfers effectively to challenging paediatric ECG interpretation tasks

Paediatric ECGs differ substantially from adult recordings in waveform morphology, heart rate distribution and developmental electrophysiological characteristics. ZZU-pECG allows assessing transferability to these settings. Despite distributional differences, HuBERT-ECG achieves robust performance (Fig. 4d), with AUROCs ranging from 0.865 (LARGE) to 0.877 (SMALL). On ZZU-pECG, HuBERT-ECG surpasses KED (0.865) while remaining competitive with ECGFounder (0.889). These results further support HuBERT-ECG’s representation transferability across heterogeneous populations and clinically distinct electrophysiological settings.

**Fig. 4:**
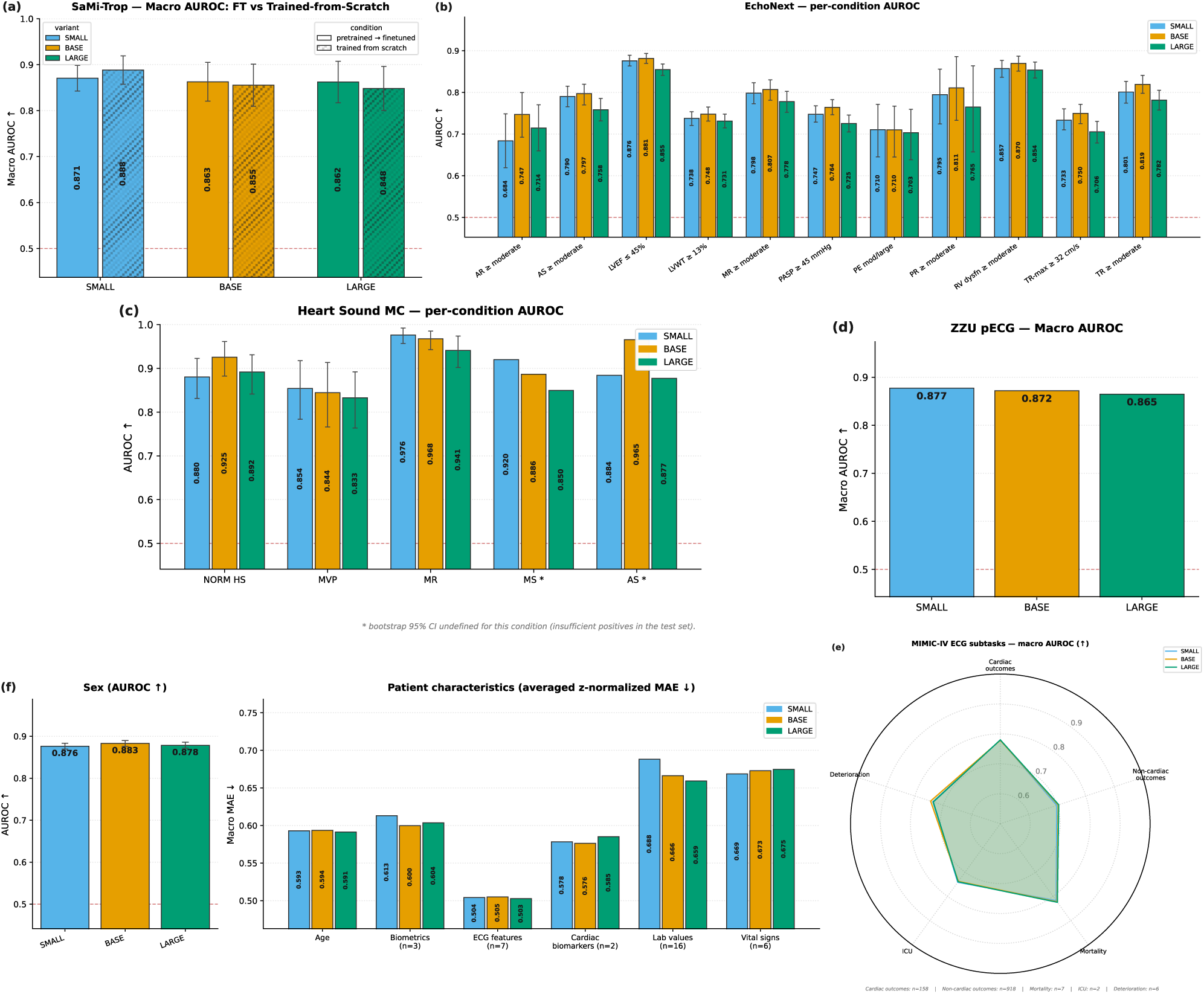
HuBERT-ECG performance on multiple highly challenging dataset comprising mostly task 2 and task 3 datasets. **(a)** HuBERT-ECG macro AUROC performance on SaMi-Trop (death prediction at 2-year follow-up); **(b)** HuBERT-ECG performance on individual type-2 tasks within EchoNext (echocardiographic findings, see Supplementary Table 2 for a description of EchoNext’s labels); **(c)** HuBERT-ECG performance on portable-device-derived single-lead ECGs from MITHSDB (type-2, heart-sound-related conditions, see Supplementary Table 3 for a description of MITHSDB classes); **(d)** HuBERT-ECG macro AUROC performance on the challenging paediatric dataset ZZU-pECG; **(f)** HuBERT-ECG performance on multiple MIMIC-IV-ECG subtasks of miscellaneous task type including sex classification, regression of age, biometrics, ECG features, cardiac biomarkers, lab values and vital signs; **(e)** HuBERT-ECG macro AUROC performance on further MIMIC-IV-ECG subtasks including cardiac and non-cardiac discharge diagnosis from the first ED recording, clinical deterioration, mortality at multiple time frames, and ICU admission. (Zoom in for better view).

##### HuBERT-ECG transfers to structural and functional cardiovascular assessment

HuBERT-ECG also transfers effectively to echocardiography-derived cardiovascular phenotyping through evaluation on EchoNext (100k ECGs, 11 tasks), a recently introduced benchmark pairing ECGs with structured echocardiographic findings (Supplementary Table 2). These findings represent prototypical type-2 task scenarios, with ECGs providing supportive electrophysiological signatures of underlying structural pathology rather than definitive diagnoses. Across these intrinsically complex echocardiography-linked tasks, fine-tuned HuBERT-ECG achieves strong per-task AUROCs ranging from 0.684 to 0.881 (Fig. 4b), with BASE performing best (macro-AUROC=0.793±0.029). Columbia-mini^53^ is the state-of-the-art on EchoNext (macro AUROC=83%) thanks to joint training with ECG traces and echocardiography features. Our performance demonstrates transferability beyond ECG-defined diagnostic findings and shows that HuBERT-ECG representations learned *solely* from ECG signals capture clinically meaningful information associated with structural and functional cardiac abnormalities identifiable through imaging.

#### HuBERT-ECG transfers to future CVE, prognostic, and acute-care prediction tasks

Predicting future CVEs (type-3 task) is exceptionally challenging. SaMi-Trop^52^ and MIMIC-IV-ECG enable assessing HuBERT-ECG’s predictive capabilities in prognostic and acute-care settings.

On SaMi-Trop (1600 ECGs from Chagas patients, 104 mortality outcomes, no censoring) HuBERT-ECG performs best with SMALL outperforming BASE and LARGE (AUROC: 0.871±0.028 vs 0.863±0.042 and 0.862±0.045; Fig. 4a). The limited event count makes results highly sensitive to misclassifications. Compared to Ferreira et al.^61^, who combined engineered ECG features, sociodemographic variables and self-reported symptoms to fit a Random Forest, HuBERT-ECG uses *only* ECG traces yet achieves superior performance under matched metrics and thresholds (*G*_*mean*_=0.78 vs 0.77). AUROC is not directly comparable due to missing model outputs and code. When fine-tuned, ECGFounder performs substantially worse and unstable (AUROC=0.702±0.348) while KED does not surpass near-random-chance scores, suggesting poor robustness in low-event regimes.

We further evaluate HuBERT-ECG on MIMIC-IV-ECG, which includes ED ECG-based prediction tasks covering cardiac outcomes, non-cardiac discharge diagnoses, clinical deterioration, ICU admission, and multi-horizon mortality (Fig. 4e). Across endpoints, all HuBERT-ECG variants perform similarly, with AUROCs of 0.779±0.050–0.780±0.049 (cardiac outcomes), 0.698±0.058–0.705±0.057 (non-cardiac outcomes), 0.737±0.073–0.745±0.090 (deterioration), 0.738±0.016–0.742±0.017 (ICU admission), and 0.818±0.032–0.825±0.035 (mortality). Compared with ECGFounder and KED, HuBERT-ECG remains competitive overall, consistently outperforming KED and matching ECGFounder on key endpoints, particularly mortality and non-cardiac outcome prediction (Table 3).

**Table 3.**
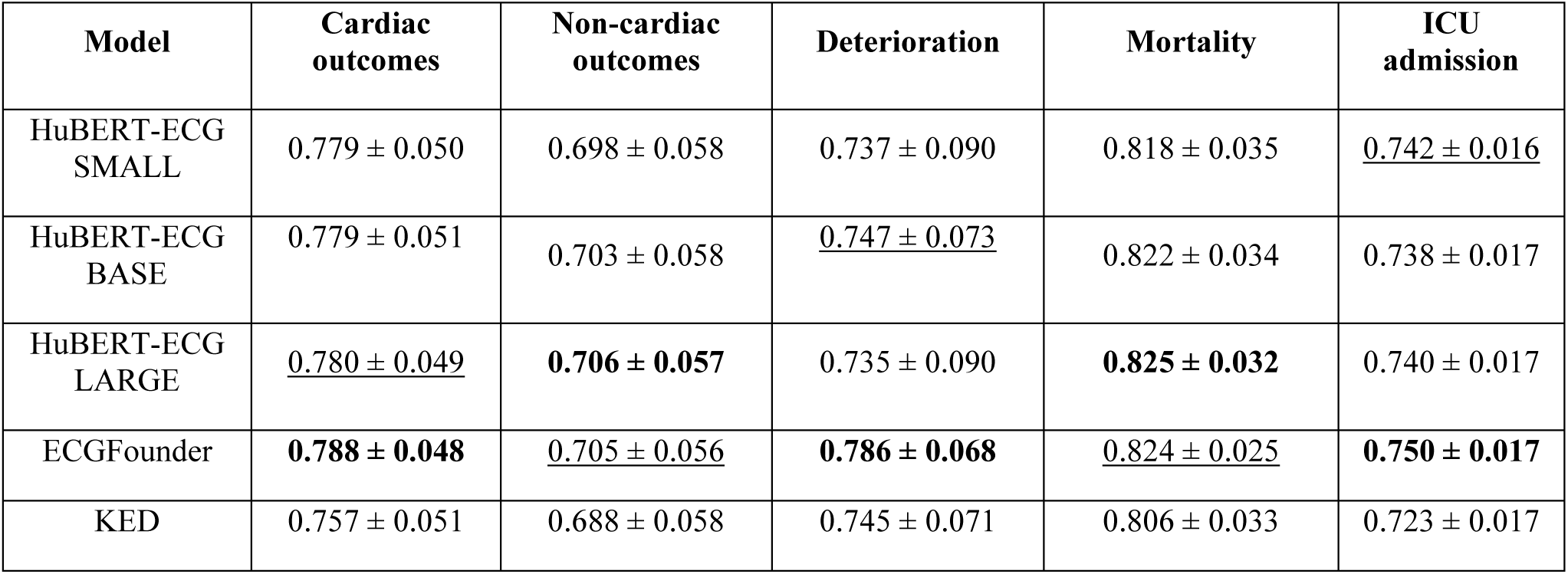
Macro AUROC performance of HuBERT-ECG and other foundation ECG models on MIMIC-IV-ECG prognostic and acute-care prediction tasks. Uncertainty is reported as 95% confidence interval. Best results are reported in bold; second-best results are underlined.

##### HuBERT-ECG captures demographic and physiological information from ECGs

We also evaluate whether HuBERT-ECG representations encode broader patient-level signals using MIMIC-IV-ECG-derived tasks spanning demographic inference, biomarker estimation, ECG feature reconstruction, laboratory values and vital sign regression (Fig. 4f, Table 4). Across heterogeneous targets, HuBERT-ECG achieves consistent performance: sex prediction AUROC up to 0.883, age MAE∼0.59, biometrics MAE∼0.60, ECG feature reconstruction MAE∼0.50, lab values MAE∼0.65, and vital signs MAE∼0.66. All variants behave similarly, with BASE typically slightly stronger or on par with SMALL and LARGE. HuBERT-ECG consistently outperforms KED across tasks and remains competitive with ECGFounder, which shows stronger sex classification but is matched or approached on most regression tasks (ECG features, lab values, vital signs). These results indicate that HuBERT-ECG’s representations encode patient-level physiological information beyond standard rhythm and diagnostic signals.

**Table 4.**
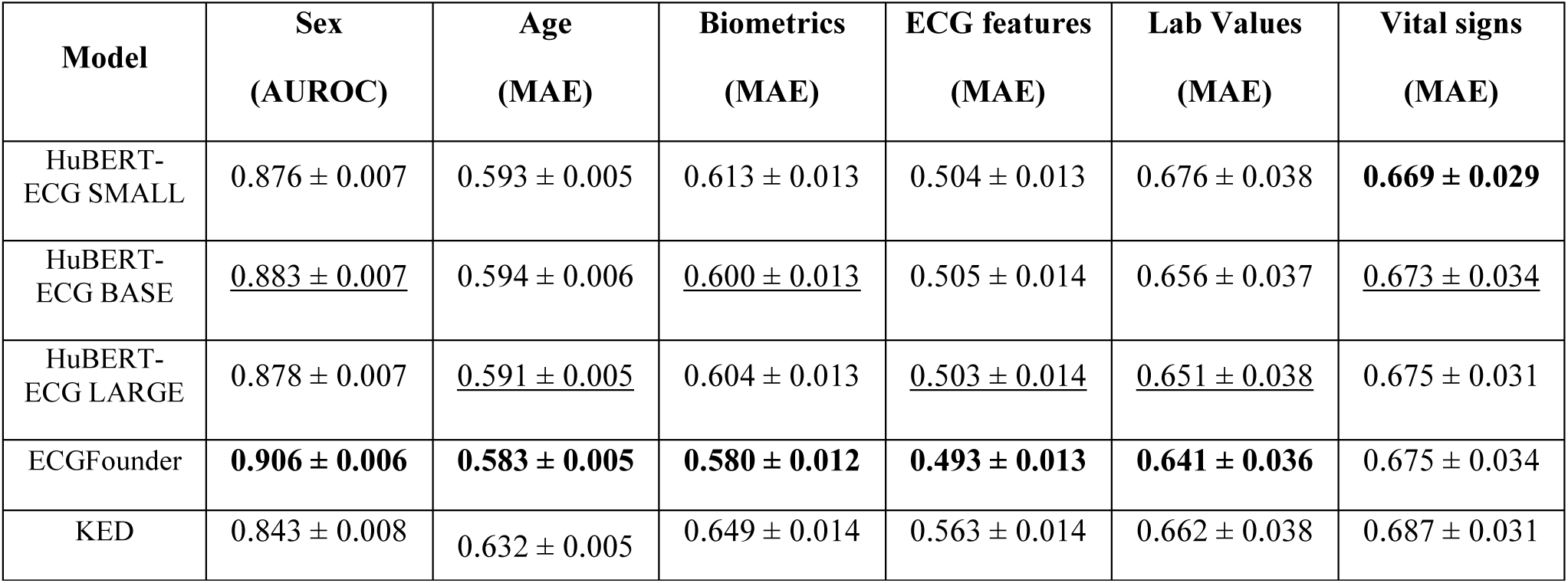
Performance in terms of AUROC and MAE of HuBERT-ECG and other foundation ECG models on patient characteristic prediction tasks. Uncertainty is reported as 95% confidence interval. Best results are reported in bold; second-best results are underlined.

##### HuBERT-ECG transfers to single-lead ECG analysis

12-lead ECGs are the clinical standard but signals with fewer derivations are increasingly recorded. Employing 12-lead models on these recordings represents a significant drift requiring substantial retraining. To assess HuBERT-ECG’s transferability beyond standard acquisition settings, we utilize the bi-modal MITHSDB using *exclusively* single-lead ECG recordings (Fig. 4c). Despite pre-training on 12 leads *only*, HuBERT-ECG shows strong per-class performance and macro AUROCs (0.878-0.917). We outperform KED (0.896) but lag behind ECGFounder (0.965). When considering the abnormality detection task proposed by MITHSDB’s authors^56^, we surpass both (0.966 vs 0.957 and 0.959, respectively), demonstrating overall transferability to portable-device-like single-lead ECG settings.

#### HuBERT-ECG navigates large-scale multi-task finetuning and improves upon task-specific training

To emulate real-world cardiovascular practice (overlapping cardiovascular conditions and heterogeneous prediction objectives) and explore benefits of multitask learning beyond isolated task-specific training, we perform large-scale multitask fine-tuning on Cardio-Learning, a new harmonised benchmark integrating 2.4 million ECGs and 164 cardiovascular tasks spanning ECG-defined findings, supportive diagnostic tasks and prognostic outcomes. Fine-tuning consistently outperforms training from scratch and HuBERT-ECG BASE tops over SMALL and LARGE sizes (AUROC=0.852±0.019 vs 0.845±0.006, 0.840±0.038) (Fig. 5a-c). When tasks are stratified by type, HuBERT-ECG BASE performs best on type-1 (0.847±0.111) and type-2 (0.867±0.058) tasks. Most interestingly, without additional recordings beyond those already present, this multitask finetuning protocol allows HuBERT-ECG BASE to raise performance on future death prediction (type-3) to 0.910±0.078 (Fig. 6a), significantly surpassing itself when fine-tuned solely on SaMi-Trop (Fig. 4a). Relevantly, when conducting multitask finetuning on Cardio-Learning, we observe the same performance increment without additional supervision even for other clinically relevant type-2 tasks (Fig. 6b).

**Fig. 5.**
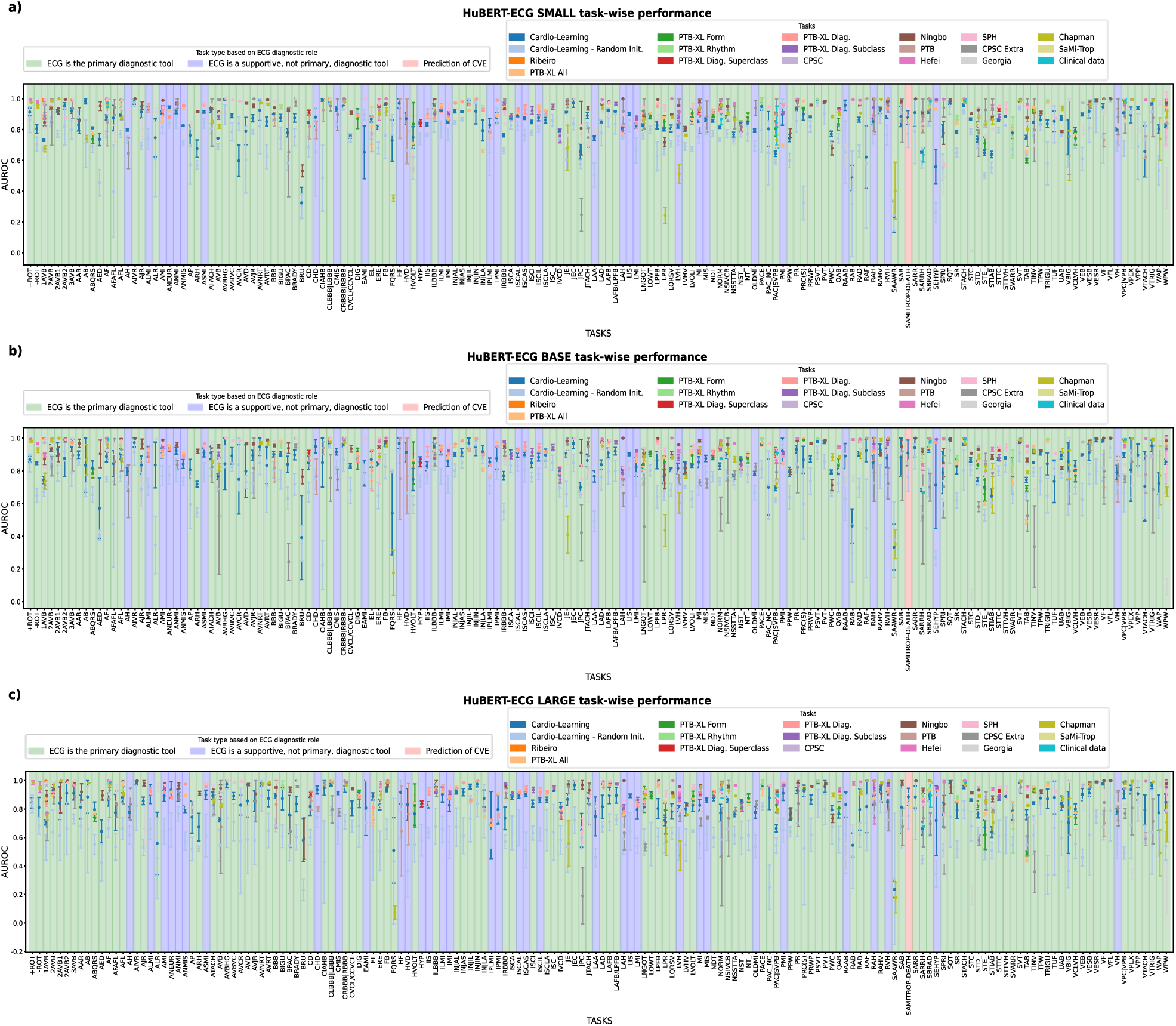
HuBERT-ECG label-wise performance for **(a)** SMALL, **(b)** BASE, and **(c)** LARGE configurations across all datasets, including Cardio-Learning. The reported AUROC values are averages obtained from cross-validation and are presented with standard deviations if a label comes a dataset with no predefined test set. Otherwise, if a label comes from a dataset with a predefined test set, uncertainty is expressed with 95% confidence intervals obtained through empirical bootstrapping using 1000 bootstrap iterations. Label backgrounds are coloured based on the diagnostic role of the ECG. Supplementary Table 1 provides label abbreviations and their corresponding diagnoses. (Zoom in for better view).

**Fig. 6.**
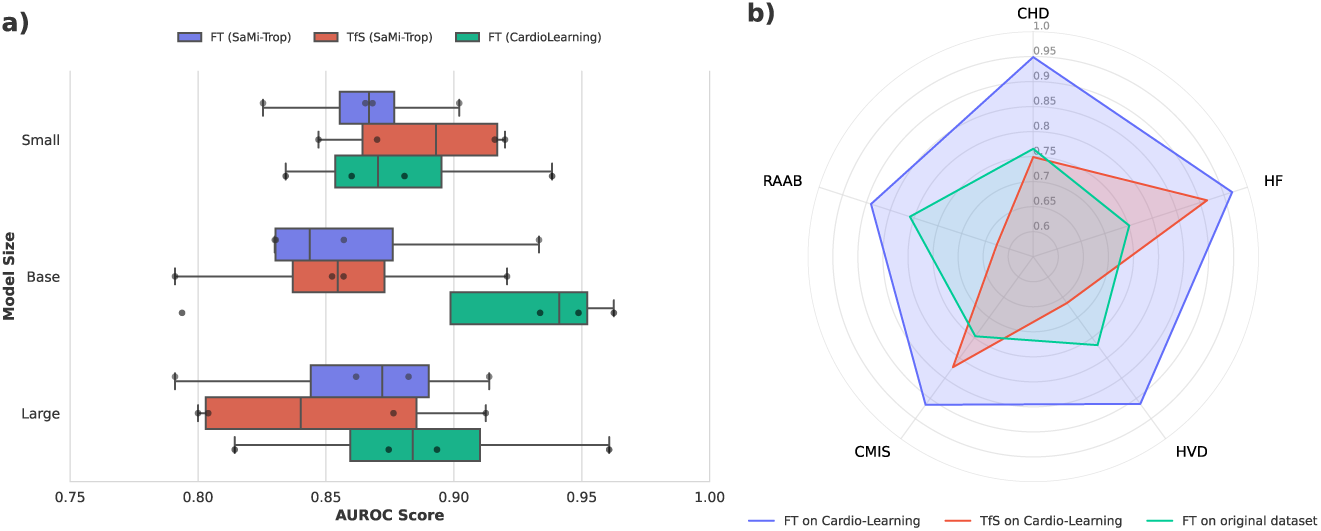
HuBERT-ECG’s performance overview with focus on diagnostic accuracy for conditions for which the ECG is used as an informative support of for predicting future CVEs. **(a)** HuBERT-ECG AUROC trend in the prediction of death events for SaMi-Trop patients when it is fine-tuned on Cardio-Learning, trained from scratch on Sami-Trop, and fine-tuned on SaMi-Trop; **(b)** HuBERT-ECG averaged AUROC performance for conditions for which the ECG is an informative support for diagnosis and that appear only in one downstream dataset when the model is (1) fine-tuned on Cardio-Learning, (2) trained from scratch on Cardio-Learning, and (3) fine-tuned on the original downstream dataset. “FT” means fine-tuning; “TfS” means training from scratch. Conditions abbreviations: “HF” means Heart Failure; “CHD” means Coronary Heart Disease; “RAAB” means Right Atrial Abnormality; “CMIS” means Chronic Myocardial Ischemia; “HVD” means Heart Valve Disorder. (Zoom in for better view).

#### HuBERT-ECG surpasses reported human performance

To assess clinical relevance, we retrospectively compare fine-tuned HuBERT-ECG on the Ribeiro dataset against human readers from Ribeiro et al.^44^, including two 4th-year cardiology residents, two 3rd-year emergency residents, and two 5th-year medical students whose paired reported sensitivity and specificity scores were averaged across labels. Using balanced accuracy as aggregated metric and discriminator (Table 5), always at least two HuBERT-ECG configurations outperform human readers on every condition, and all configurations always surpass at least two reader groups. Importantly, we note that this is not a strict head-to-head comparison, and no statistical testing is possible to missing individual human predictions. Nevertheless, results suggest fine-tuned HuBERT-ECG can match or exceed human-level performance on key diagnostic tasks, supporting its potential clinical utility.

**Table 5.**
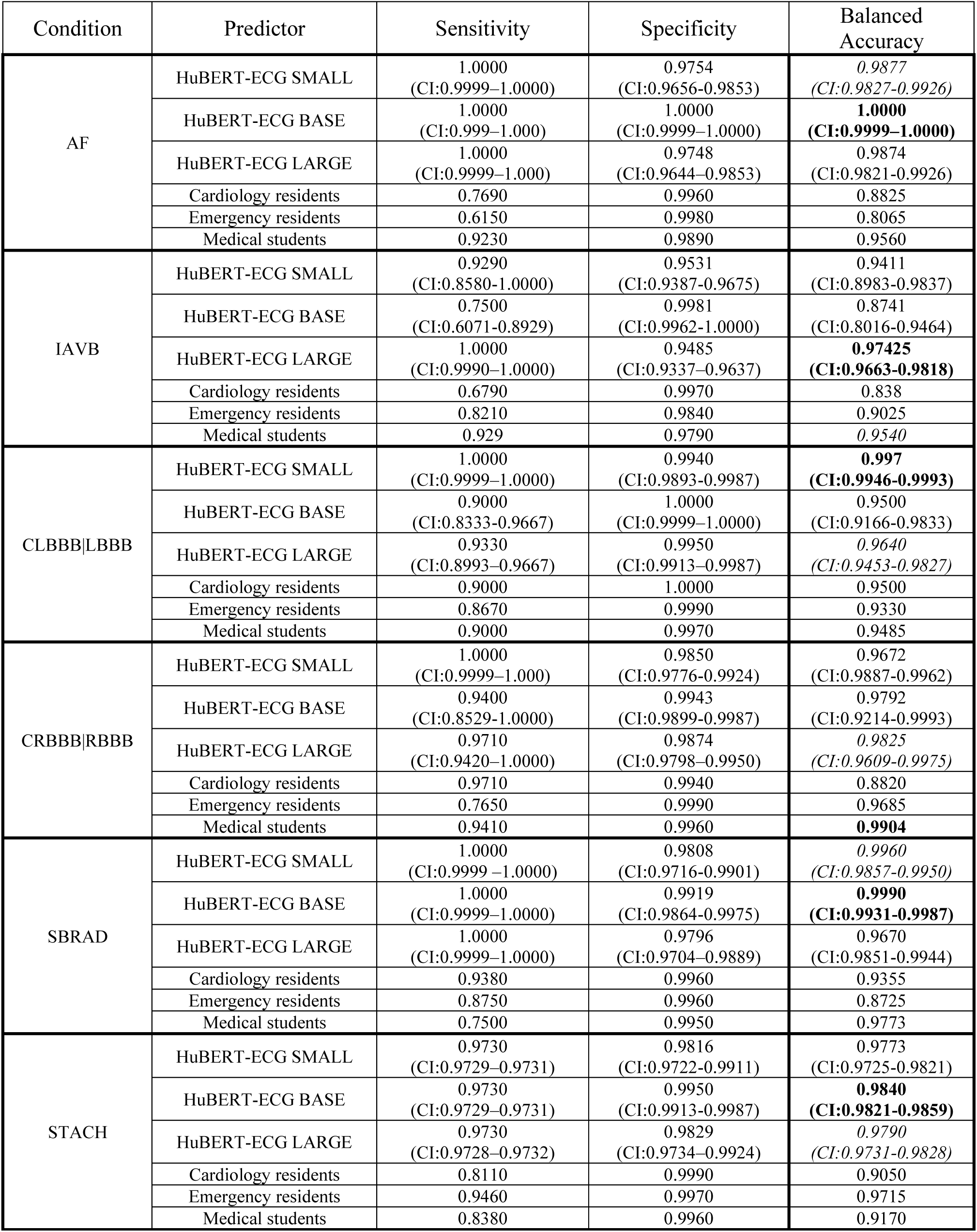
Fine-tuned HuBERT-ECG performance on Ribeiro benchmarked against two 4^th^ year cardiology residents (Cardiology residents), two 3^rd^ year emergency residents (Emergency residents), two 5^th^ year medical students (Medical students) involved in the study by Ribeiro et al. ^44^. Both residents’ and students’ performance are averages, as reported in the original study. Label abbreviations and corresponding diagnoses can be found in Supplementary Table 1.

##### Fine-tuning HuBERT-ECG is substantially more efficient than training from scratch

Beyond data labelling bottlenecks, compute cost is a major constraint in deep learning pipelines due to extensive training, tuning, and ablation requirements. As shown in Table 6, pre-trained HuBERT-ECG significantly reduces computation across settings, since self-supervised pre-training already covers expensive optimization steps that would otherwise be repeated. Even in the most demanding case—LARGE on Cardio-Learning—the full fine-tuning pipeline completes in less than 2.5 hours on a single NVIDIA A100 GPU (less than 1 kWh total energy), indicating low environmental cost. Resource usage remains modest: ∼1 GB GPU memory (up to ∼1.6 GB with fine-tuning batch size of 64), and inference latency of 34±13 ms on consumer-grade platforms such as Google Colab. Overall, HuBERT-ECG provides a strong efficiency–performance trade-off, enabling deployment and experimentation without high computational barriers.

**Table 6.**
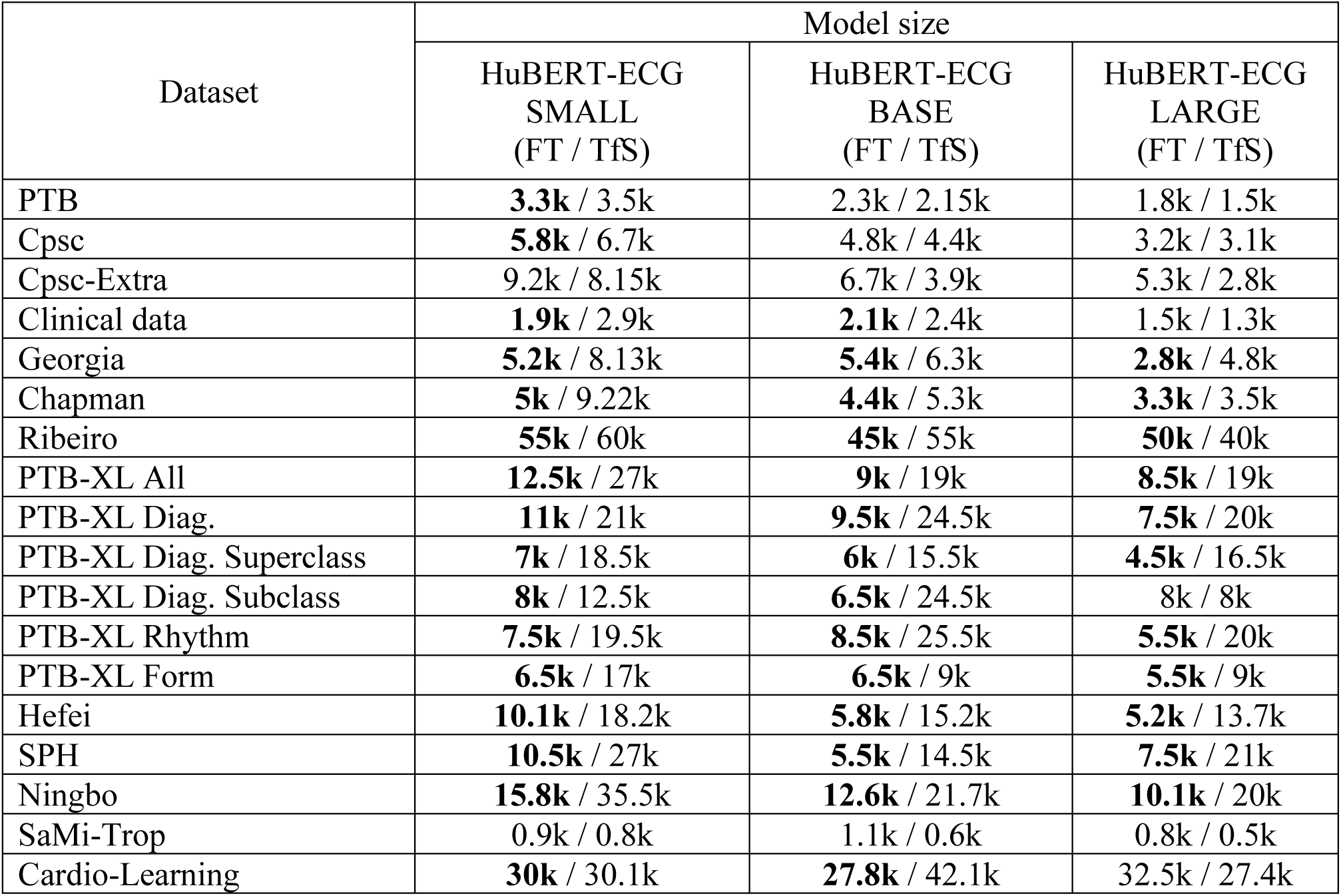
Comparison between fine-tuning (FT) and training-from-scratch time (TfS), on average, across datasets and model sizes. Time is expressed as number of training steps.

## Discussion

We present HuBERT-ECG, a self-supervised ECG foundation model pre-trained on 9.1 million ECGs collected across four countries and evaluated extensively through fine-tuning on 21 independent datasets spanning diagnostic, structural, functional, prognostic, paediatric, acute-care and single-lead acquisition settings. Collectively, our results show that our large-scale self-supervised ECG representation learning can generalise across many clinically heterogeneous tasks substantially beyond conventional rhythm and conduction analysis. These findings suggest that HuBERT-ECG’s representations encode transferable cardiovascular information not only for conditions directly observable from the tracing itself, but also for structural abnormalities, systemic physiological states and future clinical outcomes indirectly associated with ECGs.

Central in our study is evaluating systematically the transferability across clinically distinct task abstractions. We group downstream tasks into three broad categories: type-1, where ECGs are gold-standard diagnostic tools; type-2, where ECGs provide supportive but non-definitive diagnostic information; type-3, where ECGs provide prognostic signals. Unsurprisingly, performance decreases from type-1 to type-2 and type-3 tasks, reflecting increasing indirectness of underlying targets. Nevertheless, HuBERT-ECG consistently demonstrates strong transferability across all three categories, suggesting that its representations encode latent electrophysiological information shared across cardiovascular phenotypes, including conditions typically requiring imaging, biomarkers or longitudinal follow-up for definitive diagnosis.

The strong performance observed on structural and functional cardiovascular assessment tasks further supports this interpretation. In particular, HuBERT-ECG demonstrates robust transferability to echocardiography-derived targets without access to imaging data during pre-training or fine-tuning. This finding suggests that HuBERT-ECG’s representations encode clinically meaningful correlates of cardiac structure and function that can later be associated with imaging-derived phenotypes through downstream supervision. Similarly, transferability to acute-care prediction, laboratory estimation, demographic inference and physiological regression tasks indicates that its representations capture broader systemic and patient-level information beyond canonical ECG interpretation.

Another major finding concerns large-scale multitask fine-tuning on Cardio-Learning. Jointly optimising HuBERT-ECG across up to 164 cardiovascular tasks leads to substantial improvements for several challenging, low-prevalence, or weakly supervised diagnostic and prognostic targets. These gains suggest that cardiovascular tasks share non-trivial exploitable relationships that can be captured through joint optimisation. Importantly, no additional ECG is introduced beyond the original dataset: gains emerge from shared supervision across heterogeneous phenotypes. Beyond improving performance, these results support multitask learning as a practical strategy for future users for addressing sparse, imbalanced or weakly supervised ECG problems frequently encountered in real-world task-specific datasets.

We additionally analyse how performance scales with model and dataset size. While larger architectures generally benefit from large and heterogeneous datasets, scaling is not uniformly beneficial across settings. In small or highly imbalanced datasets, SMALL and BASE often outperform LARGE, indicating that more parameters alone do not guarantee better transfer in ECG representation learning. In contrast, LARGE models are more advantageous in data-rich and heterogeneous multitask scenarios, where complexity can be better modelled. Overall, the relationship between model scale, data regime, and transferability is more nuanced than the monotonic scaling trends typically observed in vision or language foundation models. These findings, combined with efficient fine-tuning, provide practical guidance for future users of HuBERT-ECG.

HuBERT-ECG’s broad transferability across heterogeneous populations and acquisition settings further supports the robustness of its representations. Despite substantial electrophysiological differences between adult and paediatric ECGs, HuBERT-ECG transfers effectively to paediatric ECG analysis tasks. Similarly, strong performance on single-lead ECG benchmarks demonstrates that representations learned only from standard 12-lead recordings retain meaningful transferability even under reduced spatial information and portable acquisition settings. Together, these findings suggest that HuBERT-ECG’s representations capture stable electrophysiological structure that generalises across demographic groups, recording conditions and device configurations.

Importantly, our findings should not be interpreted as evidence that ECG alone can replace established diagnostic modalities (e.g., echocardiography, laboratory testing, longitudinal clinical assessment). Instead, our results indicate that (i) ECG signals contain richer information than traditionally exploited in routine interpretation; (ii) our self-supervision can extract transferable representations associated with multiple clinically relevant phenotypes. HuBERT-ECG may therefore serve as computational backbone capable of supporting a broad range of downstream diagnostic and predictive applications while complementing, rather than replacing, existing workflows.

Nevertheless, limitations should be acknowledged. First, although our pre-training corpus spans multiple countries and populations, important geographic regions remain underrepresented (e.g., Africa, India). Expanding pre-training diversity will likely reduce any population-specific bias. Second, publicly available datasets for type-2 and type-3 tasks remain comparatively scarce and often smaller than datasets focused on type-1 tasks. This limitation restricts evaluation breadth. Third, despite careful harmonisation, many publicly available ECG datasets inevitably contain varying degrees of label uncertainty arising from differences in annotation protocols, physician interpretation and retrospective label extraction procedures. Such variability affects the entire ECG-AI field and likely imposes an upper bound on achievable performance for several downstream tasks. Finally, direct benchmarking against prior systems remains difficult because many studies evaluate only restricted subsets of available labels, use non-standard protocols or do not release code and predictions, thereby limiting strict reproducibility and fair comparison.

Overall, our findings support the emerging view that large-scale self-supervised learning can transform electrocardiography from a collection of specialised predictive models into a more generalisable representation-learning framework. By demonstrating robust transferability across diagnostic, structural, prognostic, physiological and demographic tasks, HuBERT-ECG presents itself as a broadly informative cardiovascular backbone applicable across diverse clinical scenarios. We hope releasing pretrained models and code will facilitate future cardiovascular research.

## Methods

### Data and Preprocessing

While many ECG-focused studies addressing general clinical questions primarily rely on datasets from the Physionet Challenges^62,63^, overlooking other substantial and valid sources, HuBERT-ECG self-supervised pre-training leverages both public and access-on-demand 12-lead ECG datasets. These include the labelled and unlabelled partitions from Ribeiro^44^ (except the predefined, publicly available test-set), CPSC and CPSC-Extra^45^, PTB^46^ and PTB-XL^47^, the publicly available partition from Georgia^48^, Chapman-Shaoxing^49^ and Ningbo First Hospital^50^, the partition from the Tianchi Arrhythmia Competition^b^, Shandong Provincial Hospital^51^, and MIMIC-IV ECG^55^. For supervised fine-tuning and evaluation we reuse all the aforementioned sources and include the SaMi-Trop^52^ dataset, the so called “Clinical data” from the work of Tian et al.^41^, EchoNext ^53^, MITHSDB^56^, and the challenging paediatric ZZU-pECG dataset ^64^ Noteworthy from a clinical point-of-view,EchoNext addresses multiple predictions tasks related to echocardiographic findings on cardiac structure and function; MIMIC-IV spans many tasks including cardiac and non-cardiac discharge diagnoses from the first emergency department (ED) ECG, acute care predictions (clinical deterioration, mortality at multiple time points, Intensive Care Unit (ICU) admission), and patient characteristic prediction (age, sex, biometrics, ECG features, lab values, and vital signs); ultimately, MITHSDB, which natively comes as bimodal dataset and incorporates portable device-derived phonocardiograms and single-lead ECGs, tackles heart sound detection using single-lead recordings, thus representing a significant distribution drift w.r.t. what HuBERT-ECG sees during pre-training. We provide a summary of the datasets used in this study in Table 7. We instead exclude the unlabelled partition of Ribeiro. Although some datasets appear during both self-supervised pre-training and supervised fine-tuning, this a widely accepted practice in developing foundation models that undergo a self-supervised pre-training. Notable examples include those by Caron et al.^21,65^ in computer vision, Hsu et al.^24^ and Baevski et al.^23^ in speech representation learning, Azizi et al.^66^, Pai et al.^67^ and Haghighi et al.^68^ in various types of medical image analyses, Ritcher et al.^69^ in cell genomics, and ultimately Mehary & Strodthoff^38^ and Tang et al.^70^ in the ECG domain. In addition, a recent survey^71^ on the impact of self-supervised pre-training for diagnostic tasks involving many different types of medical data, points out that pre-training with self-supervision and fine-tuning a model on the same data brings no tangible benefit. This is due to the lack of any information leakage, as no clinical/downstream labels or task-specific signal is exposed during the self-supervised pre-training. In our case, pre-training target variables are also pseudo-labels, discovered in an unsupervised manner using feature descriptors (MFCCs and latent representations) that do not carry any disease-specific, clinical, or dataset-specific information. For self-supervised pre-training, all labels and clinical annotations are removed from the pre-training datasets, resulting in an unlabelled dataset of 9.1 million ECGs (Fig. 2a–Pretrain). The effectiveness of the data selection in capturing the diversity of ECG signals is illustrated through a UMAP^72^ projection of the ECG embeddings (Fig. 1e). To assess HuBERT-ECG’s downstream utility on clinical tasks and datasets of varying complexity, we fine-tune it on each collected dataset^c^ and test it on hold-out partitions of such data sources that were not included in the pre-training phase. Additionally, we create a challenging new dataset, which we name *Cardio-Learning*, by merging all the above sources (except “Clinical data”, MIMIC-IV, EchoNext, Heart Sound, and ZZU-pECG) into a new one comprising over 2.4 million ECGs with 164 potentially co-occurring conditions. These conditions can be grouped into three categories based on the diagnostic role of the ECG, as shown in Fig. 1f. During the creation of Cardio-Learning, to ensure label consistency across different sources, diagnostic labels are harmonised independently by three expert cardiologists. First, diagnostic labels that appear under slightly varying nomenclature are consolidated using unique terms. For example, all instances of atrial fibrillation are uniformly labelled as “AF” instead of “AFIB” or “AF_” (see Supplementary Table 1). Second, diagnostic concepts that appear under multiple synonyms are standardised to a single diagnostic category. For instance, following established practices^62,63^, ECGs labelled as premature atrial contraction (PAC) or supraventricular premature beats (SVPB) are standardised to PAC|SVPB, as both conditions refer to the same concept. Third, hierarchically broader diagnostic categories are assigned to ECGs already labelled with more specific ones (e.g., Left Bundle Branch Block ECGs also receives the Bundle Branch Block label). However, we avoid two actions: (1) we do not infer more specific conditions from more general labels unless explicitly confirmed in the original annotation (e.g., Bundle Branch Block ECGs do not receive the Left Bundle Branch Block label); (2) if a constituent dataset includes both a more specific and a more general condition, but explicitly does not assign the general condition to ECGs labelled with the specific one, we respect this distinction and do not add it. “Clinical data”, MIMIC-IV, EchoNext, Heart Sound, and ZZU-pECG have been excluded from the creation of Cardio-Learning due to late acquisition and labels impossible to harmonize with adequate confidence.

**Table 7.**
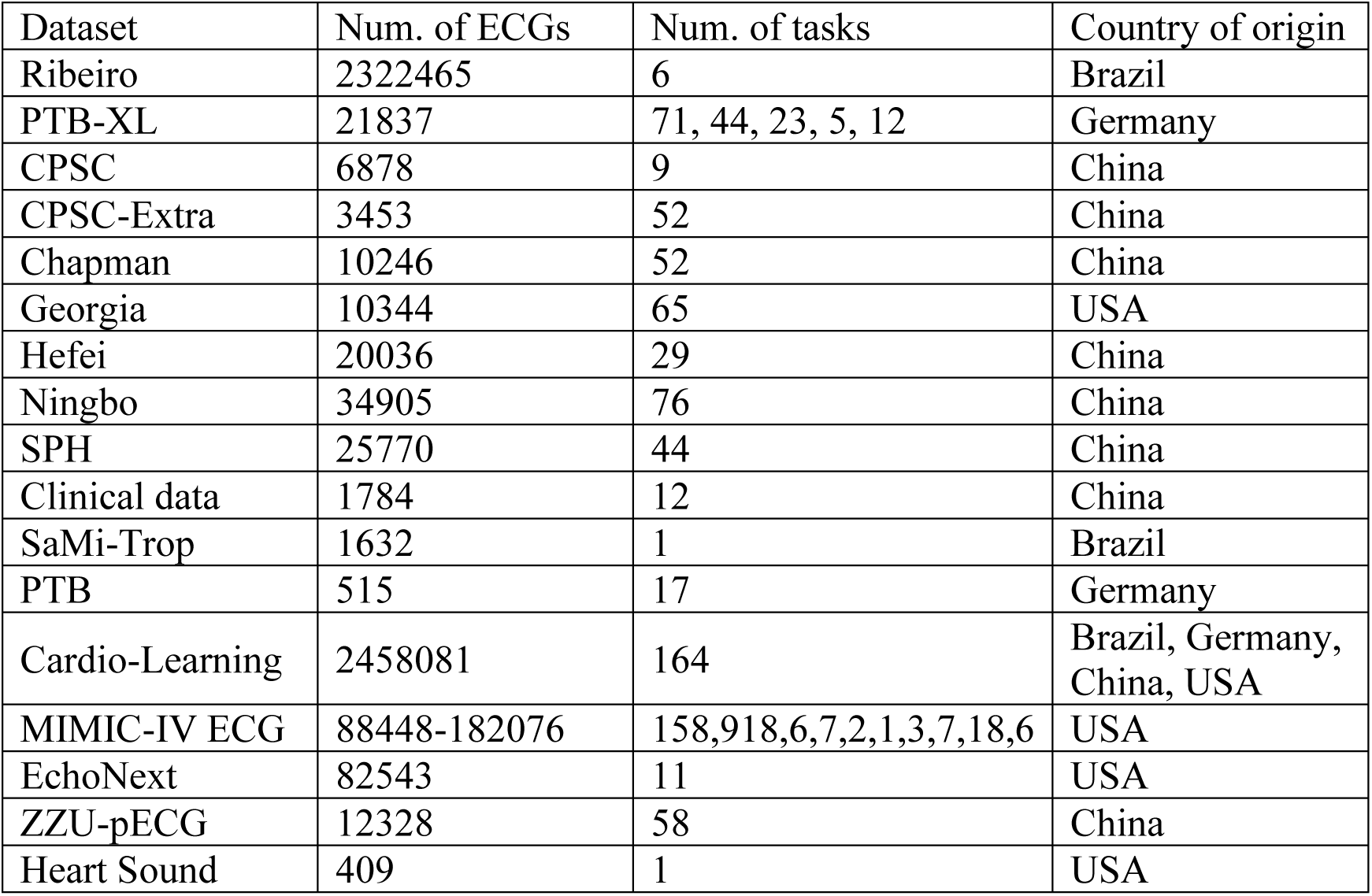
Summary of the datasets used in this study.

Remarkably, as shown in Figs. 1b and 1d, the ECGs used in this study were obtained from patients exhibiting a broad age distribution and diverse geographical origins spanning four countries. To the best of our knowledge, this constitutes one of the largest and most heterogenous dataset ever assembled to date in terms of the number of conditions, demographics, and geographic origin of the individuals.

For pre-processing, consistent with the approach of Natarajan et al.^73^, a finite impulse response bandpass filter is applied to exclude frequencies outside the [0.05, 47] Hz range, which is reported to encompass the dominant components of P waves, T waves and QRS complexes^74^. Subsequently, we investigate the impact of the sampling rate, which has no standard value and regulates the degree of dilution of the information content, on both the upstream and downstream performance. Our experimental findings indicate that resampling the ECGs at 100 Hz provides a suitable trade-off between downstream performance and training time, while preserving all the relevant physiological information content according to the Nyquist-Shannon theorem (Supplementary Information–Sec. 1.2). Following this, the ECG signal is rescaled to the [-1, 1] range, mirroring the procedure employed by Natarajan et al.^73^. Finally, unlike other works, we use 5-second 12-lead ECGs instead of 10-second recordings, hence halving memory occupation and accelerating both training and inference. In addition, the selected temporal and spectral parameters align with those derived in Mehari & Strodthoff^75^.

### HuBERT-ECG Architecture and Theoretical Framework

A schematic representation of the HuBERT-ECG architecture, its pre-training phases and its fine-tuning are shown in Fig. 2b.

#### Discovering fine-grained pseudo-labels for ECG fragments

In order to produce ground-truth pseudo-labels for the self-supervised pre-training, standard 12-lead ECGs are first flattened into one-dimensional signals and then split into non-overlapping fragments. This fragmentation is necessary to frame short portions of the ECG signal from which feature descriptors can be extracted to fit a clustering model. Its purpose is to discover and provide the fragments with cluster assignments (i.e., pseudo-labels) that are finer than the coarse and noisy ECG feature-based ones used as vocabulary for ECGBERT^76^. Conceptually, let *X* = [*x*_1_, *x*_2_, …, *x*_*N*_] be a flattened 12-lead ECG composed of *N* non-overlapping fragments *x*_*i*_, where *i* = 1, 2, … *N*. Let also *F* = [*F*_1_, *F*_2_, …, *F*_*N*_] = [[*f*_1_, *f*_2_, … *f*_*d*_]_1_, [*f*_1_, *f*_2_, … *f*_*d*_]_2_, …, [*f*_1_, *f*_2_, … *f*_*d*_]_*N*_] be the sequence of *d*-dimensional features *F*_*i*_extracted from each fragment, and *K* a clustering model fitted on *F* that finds *C* different clusters. Then, for each index *i* = 1, 2, …, *N*, under the hypothesis that *F*_*i*_ is a good descriptor of *x*_*i*_, *K*(*F*_*i*_) = *z*_*i*_ ∈ {1, 2, …, *C*} is the pseudo-label assigned to *x*_*i*_ and *Z* = [*z*_1_, *z*_2_, …, *z*_*N*_] is the sequence of these assignments for each ECG fragment. For the sake of both terminological flexibility and clarity, the terms *pseudo-labels* and *cluster assignments* will be used interchangeably.

#### Representation learning by predicting pseudo-labels for masked embeddings

HuBERT-ECG operates as a masked prediction model, henceforth denoted as *h*. When the flattened 12-lead ECG is fed into *h*, the signal is initially processed by a convolutional waveform embedder. This component captures local contextual information and generates *E* = [*e*_1_, *e*_2_, …, *e*_*N*_], a sequence of continuous ECG embeddings. Positional encodings are then integrated into the embeddings to provide the model with information about the original sequential position of each lead segment within the flattened ECG signal. This mechanism allows the model to account for inter-lead relationships that might be less evident after the 12-lead ECG is transformed into a one-dimensional sequence. Subsequently, a set of random indices *M* = {*j* | *j* ∈ {1, 2, …, *N*}}, where |*M*| < *N*, is generated to determine which embeddings are to be masked, i.e., replaced by a special learnable embedding *e*_*MASK*_. After masking, a Transformer encoder^77^, which learns global contextual information, consumes the resulting masked sequence of embeddings *E*′ and produces the output sequence of encodings *O* = [*o*_1_, *o*_2_, …, *o*_*N*_]. Eventually, a probability distribution over the *C* possible pseudo-labels is constructed for all the *N* indices: *p*_*h*_(· |*E*′, *i*) ∀*i* ∈ {1, 2, …, *N*}. This distribution is then used in a standard cross-entropy loss that, however, considers only the indices in *M*: *L*(*h*; *E*, *M*, *Z*) = ∑_*j* ∈ *M*_ *log*(*p*_*h*_(*z*_*j*_|*E*^′^, *j*))^d^. By training the model *h* to predict the cluster assignments of the masked embeddings *E*′_*j*_, ∀*j* ∈ *M*, which correspond to fragments of the input ECG not seen by the encoder, the model is forced to learn the most informative representations from the visible segments.

#### Multi-task learning via cluster ensembles

To increase the granularity of the representations being learned during the pre-training, one can use pseudo-labels generated by an ensemble of clustering models characterised by an increasing number of clusters. This approach constitutes a multi-task learning framework, where a new task is introduced with each additional clustering model. Within this framework, each embedding is associated with multiple pseudo-labels, one from each clustering model in the ensemble. The rationale behind this design is that, while a single clustering model may yield imprecise or coarse cluster assignments, an ensemble of models with an increasing number of clusters can mitigate the introduction of noisy targets and provide valuable complementary information to the model. Denoting the number of clustering models composing the ensemble as Γ, which corresponds to the number of tasks being addressed, the loss function can be reformulated as 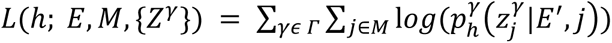, where *Z*^*γ*^ represents the sequence of cluster assignments generated by the *γ*-th clustering model and {*Z*^*γ*^} is the set of cardinality Γ comprising all such sequences.

#### Refining cluster assignments

Analogous to the work of Hsu et al.^24^, the quality of the learned representations can be further enhanced by generating more refined cluster assignments for the ECG fragments that are to be used in subsequent pre-training iterations. Importantly, this “finer” mapping does not refer to a temporal refinement, but rather to a more nuanced clustering of the features extracted from the ECG fragments. To achieve it, latent representations are extracted from intermediate layers of the partially pre-trained model *h* and then clustered. Consequently, while the cluster assignments remain aligned with the original fragments in the input *X*, their improved quality following this refinement can exert a positive impact on downstream task performance.

##### Why Transformers, HuBERT and masked modelling?

Transformers are known to excel at modelling long-range dependencies through attention, overcoming the limitations of CNNs whose fixed receptive fields restrict global context. This property underpinned the success of encoder-only NLP models such as BERT/RoBERTa ^19,78^, later adapted to speech with Wav2Vec2^23^ and HuBERT^24^. In these architectures, the tokenizer used in NLP methods to map word pieces into token embeddings is replaced by an initial convolutional module to discretize continuous real-valued signals, a necessary step given the absence of a predefined vocabulary and the continuous nature of the input signals. Despite sharing the same architecture, Wav2Vec2 was pretrained via contrastive learning, whereas HuBERT relied on masked modelling. Evidence across domains consistently suggests that masked modelling offers superior scalability and performance compared to contrastive approaches^79,80^. For these reasons, in designing HuBERT-ECG we adopt a Transformer backbone preceded by a convolutional embedder, enabling the model to capture both local morphology and global dependencies, and pre-train the resulting architecture through masked modelling to leverage its proven efficacy and scalability.

### Implementation

While the design of HuBERT-ECG mostly aligns with that proposed by Hsu et al.^24^, we implement two modifications related to the initial convolutional embedder and the masking strategy. First, since ECGs sampling rate is lower than that of audio signals, there is no need of high-stride convolutions with long filters. A shallower convolutional block with shorter filters proves equally effective. In our model, the convolutional embedder, architecturally tuned to balance embedding information granularity and computational efficiency, generates embeddings at a temporal resolution 0.64 seconds from a flattened 5-second 12-lead ECG sampled at 100 Hz, resulting in 93 embeddings. Second, instead of randomly selecting *p*% of the embeddings as starting points for masked spans, we opt to mask only the selected embeddings without spanning over adjacent ones. This approach avoids the imprecision introduced by overlapping spans, which complicated determining the exact number of masked embeddings. Our method is equivalent to constructing singleton spans and provides more accurate control over the masking process, enabling us to tune the optimal value of *p* to use during the self-supervised pre-training (Supplementary Information–Sec. 2.1). Additionally, masking individual embeddings rather than spans has also demonstrated efficacy in the work of Hu et al.^81^.

Following the masking operation, the Transformer encoder consumes the masked sequence of embeddings and produces *O*, the output sequence of encodings. The probability distribution over the cluster assignments from a generic clustering model of the ensemble is parameterised by a look-up embedding matrix *A*^*γ*^ with shape *C*^*γ*^ × *W* according to the following formula:

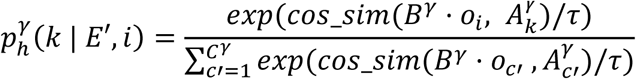

where *C*^*γ*^ denotes the number of clusters found by the *γ*-th clustering model, *B*^*γ*^ is a projection matrix used to align the dimensionality of *O* with the embedding dimension *W*, 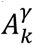 is the *k*-th row of the look-up embedding matrix *A*^*γ*^that corresponds to the *k*-th cluster assignment, *cos*_*sim*(·,·) is the cosine similarity between two vectors, and τ is the temperature that scales the logits. The superscript *γ* denotes the *γ*-th task being addressed when using an ensemble of Γ clustering models. Specifically, there are as many projection and look-up embedding matrices as there are tasks in the ensemble.

Given the extensive and heterogenous nature of our dataset, as well as the diverse needs of future fine-tuning users, we propose HuBERT-ECG in three model sizes: SMALL, BASE and LARGE. As we scale the size, we maintain the same convolutional embedder but increase the encoder depth and width, as well as the label embedding dimension *W*. Table 1 provides a detailed summary of the architectural specifications for the three model configurations. Following pre-training, to fine-tune HuBERT-ECG for specific downstream tasks, we delete the look-up embedding and projection matrices and attach a randomly initialised linear layer atop the encoder to map the pooled output sequence into logits. Specifically, we exploit the Pytorch implementation provided by Hugging Face and adapt its source code to our specific requirements. All models are trained on a computational node equipped with NVIDIA A100 GPUs. The GPU memory occupation of the largest model (i.e., HuBERT-ECG LARGE) size is approximately 1 GB and the forward pass executes in 34 ± 13 ms in freely accessible environments such as Google Colab.

### Unsupervised pseudo-label discovery

HuBERT-ECG BASE undergoes two consecutive pre-training iterations. To generate target pseudo-labels for the first one, that do not include any diagnostic or clinical information, we perform a series of k-means clustering experiments (see Supplementary Information-Sec. 1.1) and find that 100 clusters of feature descriptors consisting of 39-dimensional vectors of Mel Frequency Cepstral Coefficients^43^ (MFCCs) (13 base coefficients augmented with their first and second-order derivatives) to be a good and efficient starting point for pre-training. Furthermore, although we obtain only marginal benefit from using a cluster ensemble (see Supplementary Information–Sec. 2.2), when using multiple clusterings for pseudo-label discovery, the additional pseudo-labels are generated by two MFCC-based k-means models with 200 and 300 clusters, respectively.

For the second iteration, to produce enhanced and more refined pseudo-labels, we run the k-means algorithm again, this time increasing the number of clusters. Specifically, we use 500 clusters of latent representations extracted from the 8^th^ encoding layer of HuBERT-ECG BASE after the first iteration. Upon completion of the second iteration, we pre-train the SMALL and LARGE model configurations for one iteration using pseudo-labels generated by clustering representations extracted from 9^th^ encoding layer into 500 and 1000 clusters, respectively (Supplementary Information–Sec. 1.4). Due to memory constraints, we cannot load the entire dataset into memory and, therefore, we opt for a batched implementation of k-means wherein batches of MFCC descriptors, or latent representations, are yielded and the centroid positions are updated incrementally. We set a batch size of 9300, derived from yielding 93 descriptors from 100 ECGs, and use k-means++^82^ with 20 random restarts for a better centroid initialisation.

### Self-supervised pre-training

For each pre-training iteration we reserve 90% of the unlabelled dataset for training and the remaining 10% for internal validation. We set a batch size of 448 instances and find empirically the optimal masking percentage *p* = 33% (see Supplementary Information-Sec. 2.1) The first iteration consists of 80k training steps, while the second iteration counts 770k steps. The third iteration, instead, consists of 362.5k and 422.5k steps for the SMALL and LARGE model sizes, respectively. We use Adam optimiser with β = (0.9,0.98), an initial weight decay of 0.01 and a dropout probability of 0.1 along with a learning rate scheduler that ramps up for the first 8% of the training steps and then decays linearly to zero. Peak learning rates are set to 5e-5/5e-5/2.5e-5 for the BASE, SMALL and LARGE model configurations, respectively. In addition, we find benefits from exploiting a *dynamic regularisation* during pre-training (Supplementary Information–Sec. 1.3). This technique “penalises” the model by incrementally increasing its dropout probability and weight decay if performance does not improve on the internal validation set for *penalty-count* consecutive times. Otherwise, if performance improves on the internal validation set, with respect to the best validation loss or accuracy, the model is “rewarded” by reversing the effects of the last penalty, i.e., decreasing its dropout probability and weight decay. It is important to note that the initial weight decay and dropout probability are the minimum achievable values, while *penalty-count* emerges as a significant hyperparameter to be tuned according to the frequency with which validations are performed. During the first and third iterations, with randomly initialised models, we set the *penalty-count* to 4 and perform an internal validation every 2500 steps. For the second iteration, the internal validation is performed every 5000 steps, while maintaining the same value of *penalty-count*.

### Supervised fine-tuning

In order to assess the robustness of HuBERT-ECG’s self-supervised representations on clinically relevant datasets and tasks simulating real-world scenarios, we fine-tune the model on each labelled dataset considered in this study. These datasets differ greatly from each other in the number of instances (Fig. 1a), patients’ age distribution (Fig. 1d), and spectrum of possible conditions, each belonging to one of the three classes outlined in Fig. 1f.

When training-validation-test splits are predefined, or when at least the test set is known and defined a-priori, as in the case of PTB-XL, SPH, Ribeiro, EchoNext, ZZU-pECG, MITHSDB, or MIMIC-IV-ECG, we adhere to the original split to allow fair comparisons with previous works. However, for datasets where such a split is not available or applicable (i.e., Hefei, Ningbo, Chapman, CPSC, Georgia), we first extract a fixed hold-out test set, containing 10% of the dataset instances, using a stratified sampling to ensure that all the dataset diagnostic classes are represented. We then perform a 4-fold cross-validation on the remaining instances to tune the hyperparameters of the four models, selecting the best candidate from each fold for the evaluation on the hold-out test set. Finally, we average performance metrics that the four best candidates obtained during inference. Instead, for datasets with an extremely low cardinality compared to the model size, such as PTB, CPSC-Extra, “Clinical data”, and SaMi-Trop, we split the dataset four times in a stratified manner into four *<training, test>* folds. Then, for each fold, lacking a validation set, we skip any hyperparameter tuning and train four models until either a near-zero training error is reached or a minimum improvement delta is not achieved. We then run inference directly on the corresponding hold-out test set with the last model checkpoint. When fine-tuning on PTB, CPSC-Extra and SaMi-Trop, to prevent overfitting on these small datasets, we set a minimal early stopping’s patience of 2 to allow for transitory fluctuations in the training loss descent and set a the minimum improvement delta to 1 × 10^−3^ for both the loss function and evaluation metrics. Lastly, to construct the Cardio-Learning dataset, we merge the training, validation, and test sets from each constituent dataset to form the corresponding overall training, validation, and test partitions, ensuring the prevention of any data leakage between these partitions. We perform a 4-fold cross-validation even on Cardio-Learning, which plays simply the role of additional, challenging evaluation benchmark without constituting an intermediate learning step.

Prior to any fine-tuning, we analyse the label distribution of each dataset and exclude the instances labelled with conditions that occur only once. We do so because these conditions are either unlearnable or untestable, as their solitary instances cannot be included in both the training and test sets. However, if a condition occurs twice in the dataset, we assign one instance to the training set and one to the test set, allowing the model to attempt to learn the condition while providing a way to assess its generalisability within the limits of this setup. Eventually, when a condition occurs three times, we place one example per split. During each fine-tuning procedure, we optimise a binary cross-entropy loss function but refer to the macro-averaged multi-label AUROC on the validation set for checkpointing and model selection. The model checkpoint that yields the highest AUROC is used for inference.

To fine-tune HuBERT-ECG we follow a straightforward protocol: we attach atop the pre-trained model a randomly initialised linear layer and fine-tune all the weights of the resulting architecture, except those of the convolutional embedder. In contrast to Hsu et al.^24^, we do not use the *freezing-steps* hyperparameter, as our experiments do not demonstrate any efficacy in maintaining the encoder’s parameters fixed while training only the final linear layer. Drawing inspiration from Devlin et al.^19^, we reduce the batch size to 64 instances and the learning rate to 1e-5. Also, to gain finer control over the identification of a good model candidate, we validate our models every 50 or 500 steps, contingent on the dataset size, hence more frequently than during pre-training. Due to the high number of trainable parameters and the limited size of most datasets, HuBERT-ECG shows a tendency to overfitting but we observe no benefits from using either a strong dropout (up to 0.5) or a high weight decay (up to 0.1), nor from freezing some encoding layers. However, experiments with LayerDrop^83^ ([0.0, 0.1, 0.15, 0.2]) indicate its potential to mitigate validation loss divergence and metrics degradation. In addition, we note improvements when using a time-aligned random crop as data augmentation technique, a strategy that we also replicate at test time when we take the most confident prediction among those made on multiple crops of the same instance. This strategy helps mitigate the label-recording mismatch, which arises when beat-level phenomena (e.g., premature complexes) are used to assign labels to entire ECGs. Rather than relying on costly and far-from-perfect beat-level classifiers to locate the distinctive event, our time aligned random crop increases the chance that a label-consistent segment is framed and fed into HuBERT-ECG. At test and deployment time, this becomes certainty by covering all possible crops and selecting the most confident prediction. In summary, our findings suggest that the best performance is obtained when fine-tuning the entire Transformer encoder, zeroing the dropout probability, maintaining the weight decay at 0.01, and sweeping over the LayerDrop probability. We track all experimental runs using Weights and Biases^e^.

### Evaluation Metrics

To evaluate the downstream performance of HuBERT-ECG across the diverse datasets considered in this study, we primarily rely on the multi-label AUROC metric, which is widely used in the literature to address the challenges of multi-label classification and class imbalance^38,57,59,81^. The multi-label AUROC generalizes the concept of AUROC, originally developed for binary and multi-class problems, to multi-label scenarios where individual instances can be labelled with more than one class. In this context, the AUROC is computed independently for each condition, treating the prediction of each label as a separate binary classification task. The resulting AUROC scores for individual labels are then averaged to obtain the macro-averaged AUROC, providing a holistic perspective on the model’s performance across all diagnostic labels. The AUROC represents the area under the Receiver Operating Characteristic (ROC) curve, which graphically plots the true positive rate (TPR, or sensitivity) against the false positive rate (FPR, or 1 − specificity) at varying classification thresholds. Mathematically, TPR and FPR are defined as follows:

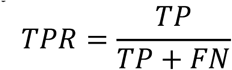

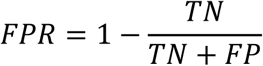

where TP, FN, TN, and FP denote true positives, false negatives, true negatives, and false positives, respectively. The AUROC value quantifies the model’s ability to discriminate between instances with or without a specific condition. A ideal classifier achieves an AUROC of 1, with a corresponding ROC curve that reaches the top-left corner of the FPR/TPR plane (coordinate (0,1)). AUROC values closer to 1.0 indicate superior model performance, while a value of 0.5 suggests no predictive capabilities no better than random chance.

On Ribeiro, the largest dataset used in our study, we are able to compute AUPRC as additional metric. This metric is based on the concept of Precision, which quantifies the proportion of correctly predicted positive instances among all instances predicted as positive for a specific diagnostic category and classification threshold. Precision is mathematically defined as follows:

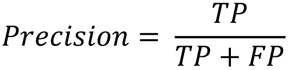

High precision values indicate that the model minimises overpredicting positive conditions, thereby reducing the occurrence of false positive alarms. The AUPRC is the area under the precision recall (i.e., sensitivity) curve and evaluates the trade-off between precision and sensitivity across varying classification thresholds. This metric reflects the model’s ability to identify true positive samples without being tricked by confounding factors into predicting false positive instances.

For regression tasks in which HuBERT-ECG has to output a real number representing, for example, the age of a subject or the value of some laboratory test, we follow standard practice and refer to Mean Absolute Error (MAE) as target metric to be minimized:

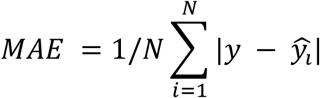

with N being the size of the evaluation set.

To guide clinically meaningful interpretation, we explicitly report expected AUROC ranges by task type, as defined in the Introduction and based on the diagnostic role of the ECG. For type-1 tasks, AUROC values between 0.9 and 1.0 are expected, as the target condition is directly encoded in the waveform. For type-2 tasks, even optimal models are inherently limited to moderate AUROC values broadly between 0.7 and 0.85, reflecting the partial information content of the signal. Finally, type-3 tasks, which aim to predict future cardiovascular events such as death or hospitalization, represent the most challenging setting. In these tasks the ECG contains only indirect prognostic markers (e.g., QT/QTc prolongation, QRS widening), so AUROC expectations are guided by prior prognostic literature on similar tasks rather than by human interpretation or expectation. To ensure comparability with prior works and following standard practices, we report dataset-level macro AUROC values (e.g. Fig.3, 4, Supplementary Table 8). In addition, to promote transparency on and inspection of HuBERT-ECG’s performance on individual conditions across all datasets, we show per-condition AUROC values as well (e.g. Fig. 5, Table 2).

## Data availability

The PTB, CPSC, CPSC-Extra, Georgia, Chapman, PTB-XL, and Ningbo datasets are publicly accessible through PhysioNet at the following URL: https://physionet.org/content/challenge-2021/1.0.3/training/ningbo/files-panel.

The Hefei dataset used in this study can be downloaded from the Tianchi platform at: https://tianchi.aliyun.com/competition/entrance/231754/information/.

The SPH dataset is available through Figshare at the following Springer Nature repository: https://springernature.figshare.com/collections/A_large-scale_multi-label_12-lead_electrocardiogram_database_with_standardized_diagnostic_statements/5779802/1.

The SaMi-Trop dataset can be downloaded from Zenodo at: https://zenodo.org/records/4905618. The “Clinical data” from the study of Tian et al. is publicly available at https://github.com/control-spiderman/ECGFM-KED.git.

MIMIC-IV-ECG dataset is available at https://physionet.org/content/mimic-iv-ecg/1.0/. EchoNext dataset is available at https://physionet.org/content/echonext/1.1.0/.

MITHSDB is available at https://physionet.org/content/challenge-2016/1.0.0/training-a/files-panel. The Ribeiro dataset is accessible for scientific research upon request to the respective owner. However, its test set is publicly available from Zenodo at: https://zenodo.org/records/3765780.

## Code availability

The full pipeline utilised in this study is available at the following GitHub repository: https://github.com/Edoar-do/HuBERT-ECG. This repository includes: (1) code for data preprocessing, from raw data management to the creation of the train-validation-test partitions used in our research; (2) scripts for replicating the pre-training (after gaining access to the pre-training dataset), fine-tuning and inference of every model developed across all datasets; and (3) code for reproducing our performance validation. Eventually, to both facilitate reproducibility and enable rapid implementation, we provide (4) pre-trained weights for all model configurations on Hugging Face^f^.

## Data Availability

All datasets supporting the findings described in this manuscript are public, except for Ribeiro. This dataset, the test set of which is publicly available, is accessible for scientific research upon request to the respective owner.

## Acknowledgements

This work has been partly funded by 1) Regione Lombardia, Italy, through the initiative “Programme of measures for economic recovery: development of new cooperation agreements with universities for research, innovation and technology transfer”-DGR n. XI/4445/2021; 2) European Union-Next Generation EU, through the Italian Ministry of Research PRIN 2022, project n. 2022A49KR3 “QT-SEED Quality-of-life Technological and Societal Exploitation of ECG Diagnostics”.

We also thank the authors of the CODE study^44^ for giving us access to the Ribeiro dataset.

## Author Contribution

**Edoardo Coppola:** Conceptualization, Data curation, Formal analysis, Investigation, Methodology, Project administration, Software, Validation, Visualization, Writing-original draft, Writing-review & editing. **Mattia Savardi:** Conceptualization, Investigation, Methodology, Project administration, Supervision, Visualization, Writing-original draft, Writing-review & editing. **Mauro Massussi:** Conceptualization, Investigation, Writing-original draft, Writing-review & editing. **Marianna Adamo:** Investigation, Writing-review & editing. **Marco Metra:** Investigation, Writing-review & editing. **Alberto Signoroni:** Conceptualization, Funding acquisition, Investigation, Project administration, Resources, Supervision, Writing-original draft, Writing-review & editing.

## Competing Interest

The authors declare no competing financial or non-financial interests.

## Ethics statement

For the Ribeiro dataset the CODE Study was approved by the Research Ethics Committee of the Universidade Federal de Minas Gerais, protocol 49368496317.7.0000.5149.

## Supplementary Information

### 1. Pre-training analyses

This section presents an analysis of how pre-training with different ECG feature descriptors and sampling rates impact downstream performance. We then investigate the interesting effects of using our dynamic regularisation during pre-training. Finally, we examine the clustering quality across both HuBERT-ECG encoding layers and pre-training iterations to measure their downstream impact. To these aims, we pre-train HuBERT-ECG BASE for one iteration with fixed hyperparameters and running configuration that are described in each of the following sections. Then, to evaluate the impact of different choices, we simply perform linear evaluations, i.e., training of a randomly initialised linear layer atop a frozen pre-trained model, on a development set that we extract from the labelled dataset used by Ribeiro et al.^1^. We build such a set, hereafter referred to as *Ribeiro-dev*, by deliberately excluding all normal ECGs, as we are more interested in assessing the ability to detect cardiac abnormalities rather than normal ECGs. Linear evaluation results represent a conservative estimate, as they are derived using fewer trainable parameters and development set that include more than 300k instances.

#### 1.1 Exploring feature descriptors for ECG segments

In order to identify the most appropriate ECG segment descriptor to use in the initial pre-training iteration, we perform k-means clustering on the entire training set with 3 different feature descriptors extracted from the ECG segments. It is important to mention that we choose k-means over other clustering approaches (e.g. DBSCAN, hierarchical clustering, Gaussian Mixture models) for its computational efficiency and scalability, two crucial ingredients when working in large-scale setups as ours. The first descriptor takes into account 16 simple time-frequency features, while the second one is based on 39 Mel Frequency Cepstral coefficients^2^ (MFCCs) following Hsu et al.^3^. These descriptors have been successfully used in ECG analysis^4,5^. The third descriptor combines the first 13 MFCCs and all the previous time-frequency features. For each descriptor, we run k-means with 10, 30, 50, 100, 150, 200 and 300 clusters and compute the corresponding sum of squared errors (SSE), or inertia (Supplementary Fig. 1). These clusters correspond to the pseudo-labels used as target variable during the self-supervised pre-training of HuBERT-ECG. We observe that MFCC- and time-frequency based descriptors provide comparable inertia and a global mean squared error in the order of 1e-5, and thus appear as promising candidates. Following the elbow method^6^, and driven by the intuition that more clusters capture too fine and specific ECG patterns for the first pre-training iteration, we set C = 100 as the best number of clusters to use irrespective of the descriptor. Given this number of clusters, we compute the Davies-Bouldin^7^ index to have an additional selection criterion and obtain extremely low scores. Interestingly, while clustering on MFCCs yields higher inertia than clustering on time-frequency features, clusters on the former are slightly more compact, as suggested by a marginally lower Davies-Bouldin score. Therefore, we experiment with the use of both MFCC- and time-frequency based k-means models during pre-training and, then, linearly evaluate the resulting models on *Ribeiro-dev* until performance plateau or a maximum number of 80k training steps is reached. The linear evaluation results, presented in Supplementary Table 2, show that pre-training with pseudo-labels generated by an MFCC-based k-means model yields better downstream performance.

#### 1.2 How the sampling rate affects downstream performance

The sampling rate of an ECG depends on the specific settings of the machine recording the electrical activity of the heart and there is no standard practice in this regard. For example, ECGs from the Ribeiro et al.^1^ dataset are sampled at 400 Hz, while ECGs from PTB-XL^8^ and SPH^9^ are sampled at 400 and 500 Hz, respectively. We investigate the effect that the ECGs sampling rate has on downstream performance. This parameter is important as it regulates the dimensionality of the data, therefore the computation speed and the memory occupation, and the degree of dilution of the information content. While it is true that training with ECGs sampled at lower frequencies is computationally less expensive, it is also true that sampling ECGs at lower frequencies may not provide enough samples to capture significant features and nuances in the input signal. Increasing the sampling rate may remedy this issue, at the cost of slower training and higher memory occupation. However, an oversampled ECG may contain redundant samples that overly dilute the information content and may be perceived as noise by a deep learning model. For these reasons, after band-pass filtering our ECGs to exclude frequencies outside [0.05, 47] Hz, which is reported to contain the dominant components of P waves, T waves and QRS complexes^10^, we investigate the effects that sampling a 12-lead ECG at 50 and 100 Hz on linear evaluation performance over *Ribeiro-dev*. For these experiments, to determine a suitable sampling rate to work with, we linearly evaluate pre-trained models until the performance plateau or 80k training steps are performed. To always have the same number of embeddings being processed by the Transformer encoder, we design waveform convolutional embedders that become increasingly shallow as the sampling rate decreases, while also maintaining the same number of ECG segments. All the other design choices and running configurations are fixed across the experiments, as shown in Supplementary Table 3. Supplementary Fig. 2a illustrates the validation loss curves observed during pre-training at 50 and 100 Hz while Supplementary Fig. 2b shows the corresponding linear evaluation performance on *Ribeiro-dev*. Two discernible trends are evident from the former: as the sampling rate decreases, the validation loss curves start at lower values than those corresponding to higher sampling rates; furthermore, as the sampling rate decreases, the models tend to show earlier overfitting to the training data. We suspect that at lower sampling rates, the information needed to solve the upstream task becomes more readily accessible, thereby facilitating the learning process. However, when clinical downstream labels are introduced, there is a noticeable performance discrepancy between models operating at 50 Hz and those operating at twice this sampling rate (Supplementary Fig. 2b and Supplementary Table 4). We attribute this discrepancy to the insufficient number of samples available to capture condition-related patterns. In summary, sampling at 100 Hz captures all the meaningful physiological information, satisfies the Nyquist-Shannon theorem, and represents a desirable trade-off between computational cost and accurate downstream performance. All results are shown in Supplementary Table 4.

#### 1.3 The effects of the dynamic regularisation

In order to gain a deeper insight into the impact of dynamic regularisation on training time and the time required to tune regularisation terms, including weight decay and dropout probabilities, we consider four different pre-training setups, each followed by a linear evaluation to assess the actual benefit on downstream performance. The first and the second ones consist in performing the entire first pre-training iteration (260k steps with no early stopping) with a weight decay of 0.01 and a dropout probability of 0.1 (referred to as *default*), with and without dynamic regularisation, respectively. The last two setups, instead, consist in performing the same pre-training iteration, with and without dynamic regularisation, but setting the initial weight decay and dropout probability to the maximum values found by the dynamic regularisation. All experiments consider the BASE architecture, ECGs sampled at 100 Hz, a MFCC-based k-means as pseudo-label generator (C = 100), a batch size of 448 instances and a masking percentage *p* = 33%. The results of our experiments, presented in Supplementary Table 5 and labelled with letters for clarity, reveal interesting insights. When using dynamic regularisation with default weight decay and dropout probability (setup A), HuBERT-ECG converges in 80k steps, resulting in the best macro-averaged AUROC of 0.933 during linear evaluation. In contrast, disabling the dynamic regularisation while maintaining the default regularisation terms (setup B), significantly slows down the convergence and leads to inferior downstream results. Instead, when comparing setups C and D, we observe two opposite behaviours: setup C shows faster convergence but worse upstream performance, while setup D shows slower convergence but better upstream performance. Despite these discordant trends, both setups produce close downstream results, both surpassing those from setup B, but still falling short of those of setup A. This suggests that pre-training with the dynamic regularisation, or the maximum regularisation terms it finds, accelerate pre-training and allows the model to adapt itself to avoid overfitting, resulting in improved linear evaluation when compared to scenarios where the dynamic regularisation is not used at all. Nevertheless, we hypothesise that initiating pre-training with already high regularisation terms, irrespective of whether dynamic regularisation is employed or not (setups C and D), may impair the model’s learning capability at the most crucial stage, resulting in inferior, albeit marginal, downstream performance. To investigate into the source of these benefits, it seems plausible that a dynamic dropout rate encourages the model to upweight alternative hidden paths in the Transformer’s feed-forward blocks when necessary, without permanently excluding the dropped ones.

In such cases, when also the weight decay increases, this complementary form of regularisation prevents significant changes to the weights of those paths, thereby maintaining training stability.

#### 1.4 The downstream impact of clustering quality across encoding layers and iterations

In the work of Hsu et al.^3^, prior to the second and third pre-training iterations, the most suitable number of clusters was determined empirically, as well as the layer from which latent features had to be extracted. This was achieved through the analysis of the quality of clusters of hidden representations extracted from each model layer. In particular, an automatic-speech-recognition model was used to produce frame-level phonetic labels that served as targets to measure the correctness of forced-aligned cluster assignments. The same approach is not applicable to our case as, to the best of our knowledge, there is no powerful open-source ECG model that can produce frame-level forced-aligned cluster assignments. Consequently, to determine which layer’s latent features should be clustered and how many clusters are to be found, we make use once again of traditional compactness and separability metrics. In addition, to limit the set of layers to explore, we combine such metrics with findings from the NLP domain regarding layers transferability^11^. After sampling 10% of pre-training ECGs, we cluster their segments’ latent representations from the 5^th^ – 10^th^ Transformer layer and measure the clustering quality in terms of inertia, Davies-Bouldin index and Calinsky-Harabasz^12^ index. We exclude from consideration shallower layers on the grounds that they would produce too coarse representations to be of use in clustering. Similarly, deeper layers are excluded on the grounds that they would generate representations that would lead to too task-specific pseudo-labels for a generic pre-training. Upon completion of the first iteration, we identify 500 and 1000 clusters of latent features from each of these layers, since we consider that setting a higher number of clusters than that used in the first iteration is necessary for two reasons: 1) to try to generate much finer cluster assignments; 2) to avoid measuring, through clustering, the degree of separability of the classes learnt during the previous iteration. The results of this final analysis are presented in Supplementary Fig. 3. As can be seen, each metric shows a monotonic trend, regardless of the representations being clustered. Therefore, they do not provide an unequivocal indication of the best encoding layer to extract features from. To continue the pre-training of the BASE configuration, we then choose to cluster latent representations from the 8^th^ layer into 500 clusters, as this point seems to mark a non-negligible change in the metrics we consider. After completing the second iteration, we re-introduce diagnostic labels and perform a linear evaluation with the same running configurations reported in previous paragraphs to quantify the relative improvement with respect to previous evaluations. As shown in Supplementary Fig. 4, when the first-iteration model saturates, the second-iteration one achieves results that are approximately 5 AUROC points better and still has room for improvement. Once HuBERT-ECG BASE completes the second iteration, we repeat the clustering step described above and report the results in Supplementary Fig. 3. Once again, the metrics we consider show a monotonic trend that does not facilitate the choice of an acceptable number of clusters (500 or 1000) nor which layer the latent representations should be extracted from. For these reasons, to start pre-training the SMALL and LARGE model configurations, we decide to extract latent features from the 9^th^ layer and to cluster them into 500 and 1000 clusters, respectively. We believe that pre-training these third-iteration models requires, for any configuration, to select a deeper layer than those selected for the previous iterations. Furthermore, increasing the quantity and fineness of cluster assignments can be beneficial for a model with higher learning capacity such as HuBERT-ECG LARGE. Conversely, for the SMALL model size, maintaining the same number and granularity of cluster assignments can facilitate the task of mimicking the BASE model without significant loss of performance. The benefits of these choices are also displayed in Supplementary Fig. 4. Upon completion of 362.5k-and 422.5k-step pre-trainings, the SMALL and LARGE configurations perform similarly when linearly evaluated on *Ribeiro-dev* for 80k steps: both achieve a macro-averaged AUROC slightly lower than that of the BASE configuration, but show no sign of strong saturation within the training time of this linear evaluation.

### 2 Ablation Study

In this section, we present a short sequence of ablation studies to investigate the effects of specific architectural choices and the impact of important hyperparameters. In particular, we study (1) the impact of our masking strategy, and (2) the effects of multi-task learning when experimenting with multiple cluster ensembles. To do so we perform multiple pre-trainings of HuBER-ECG BASE (first iteration only) followed by linear evaluations on *Ribeiro-dev* with fixed hyperparameters. If not otherwise mentioned, the experimental configurations are the same of the previous section.

#### 2.1 Impacts of the Masking Strategy

We consider setting the value of the masking percentage *p* of crucial importance to make HuBERT-ECG learn high quality representations of 12-lead ECGs. An excessively low value would generate a trivial upstream task, while an exaggeratedly high value would result into a nearly impossible one. For this reason, we experiment with multiple values of this hyperparameter in order to see how it impacts on downstream performance and plot the results of these experiments in Figure 5. For the sake of comparison, we also plot the exact percentage of embeddings that are masked when the masking strategy proposed by Hsu et al.^3^ is used. Beyond some statistical and label noise, what emerges clearly is that setting *p* = 33% guarantees the best results, as linear evaluation performance peaks at this masking percentage, while following the masking strategy used to pre-train HuBERT leads to suboptimal performance. We believe this finding is due to the more regular patterns and high information redundancy of ECGs compared to audio signals.

#### 2.2 The Effects Of Multi-Task Learning

To observe the downstream effects of pre-training HuBERT-ECG in a multi-task learning framework, we perform three pre-trainings with an increasing number of tasks to solve. Initially, we pre-train HuBERT-ECG using pseudo-labels generated by a single MFCC-based k-means model with 100 clusters. Then, we pre-train again with an additional clustering model with 200 clusters and, eventually, we consider an ensemble of three k-means models with 100, 200 and 300 clusters. Supplementary Table 6 reports the attained results. Interestingly, adding just a new clustering model does not change pre-training nor affects downstream performance. In contrast, although marginal, we see an improvement when considering an ensemble of three k-means models providing much more granular pseudo-labels. Since solving three tasks at once is harder than solving just one of them, it is not surprising to see that such performance gains follow a much longer pre-training. We also believe that the cardinality of the ensemble is not as relevant as the maximum number of clusters, which is likely more useful to the model to capture label-specific patterns in downstream data. This is analogous to the refinement of the cluster assignments that is performed prior to the second iteration, except for how such refinement is achieved, since both refinements aim to generate finer and more granular pseudo-labels for the ECG segments. Considering both the longer pre-training time and the marginal improvement obtained on the results reached after training with pseudo-labels generated by a single clustering model, we do not proceed in pre-training with cluster ensembles.

### 3 Explaining fine-tuned diagnostic models

The self-supervised pre-trained HuBERT-ECG model has no knowledge of diseases or their manifestations in ECG signals. During pretraining, the model learns only the structural and statistical properties of the signal, encoding them into general-purpose representations^13^. This learning process is guided by the pre-training task of predicting pseudo-labels—generated in an unsupervised, disease-unrelated way—for masked ECG embeddings. As such, the representations learned are not directly aligned with clinically useful, human-interpretable ECG characteristics. Because of this, applying explainability techniques directly to the pretrained model would yield explanations that are clinically uninformative: the internal features are not intended to capture pathophysiological phenomena. In contrast, once HuBERT-ECG is fine-tuned on a supervised diagnostic task, its representations become aligned with disease-specific patterns, making explainability techniques more meaningful.

In this section, we apply explainability methods to a supervised fine-tuned version of HuBERT-ECG. Specifically, we use attention maps^14^, which are well-suited to Transformer-based architectures, to visualize which parts of the ECG the model focuses on when making predictions. In Supplementary Fig. 6, we present four ECGs from the PTB-XL dataset labelled as Sinus Rhythm and correctly classified by the model. The attention maps show that the model focuses on multiple regions of the signal, with some variability across recordings. This variability is expected and desirable, reflecting inter-patient differences and signal-specific features that HuBERT-ECG learns during fine-tuning and leverages when predicting the output condition. In Supplementary Fig. 7, we show two ECGs labelled as Atrial Fibrillation, also correctly detected. In these cases, the attention maps highlight clinically relevant signal segments, suggesting that the model is focusing on meaningful characteristics rather than spurious patterns or noise.

While these visualizations do not provide a direct one-to-one mapping with classical ECG features (e.g., P-wave morphology or QRS duration), they offer qualitative evidence that the fine-tuned model attends to diagnostic signal regions in a task-consistent manner. The observed variability in attention patterns reflects the model’s flexibility in handling heterogeneous ECG presentations, rather than a lack of intelligent feature extraction.

## Supplementary Figures

**Supplementary Fig. 1.**
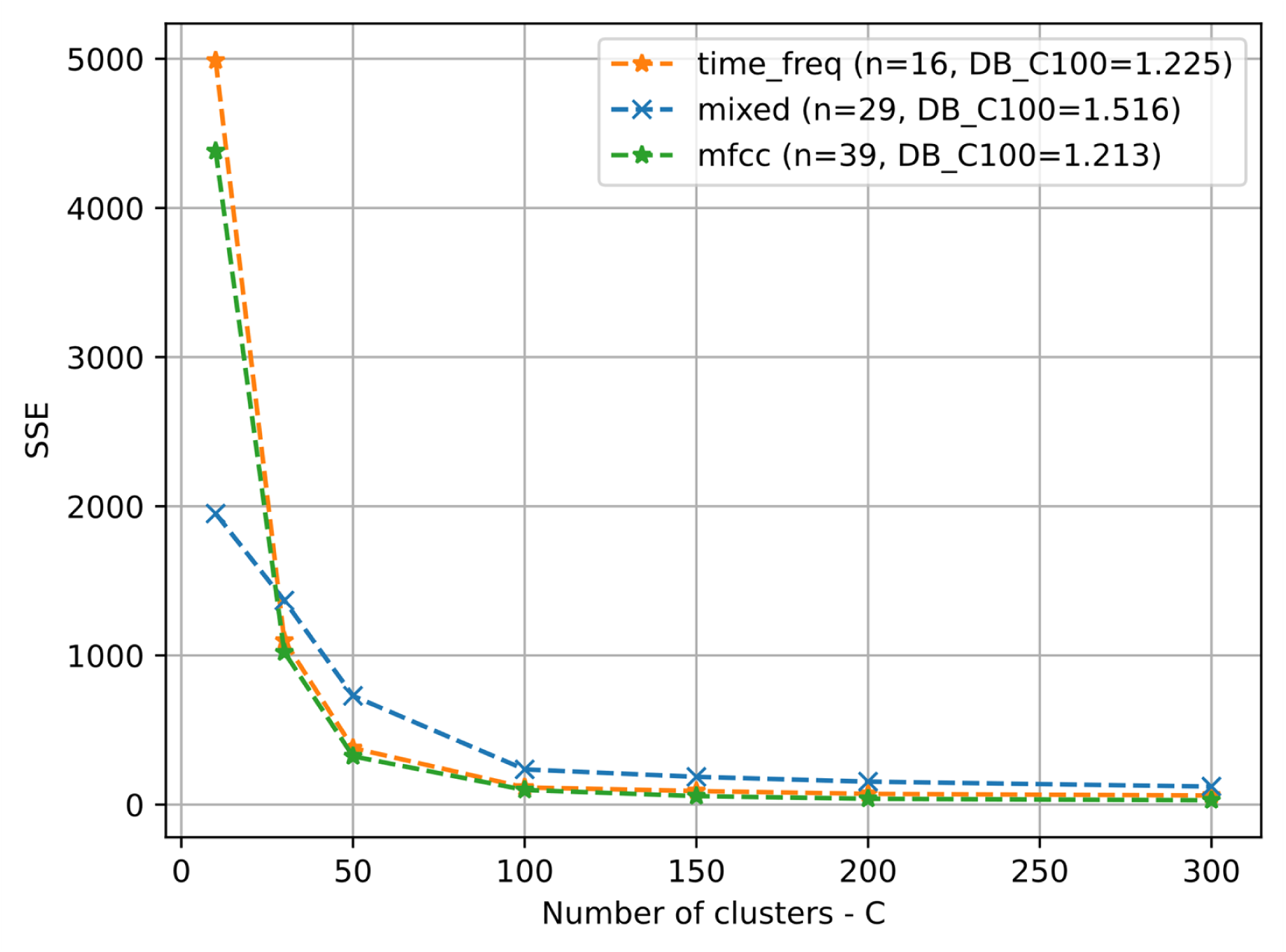
Performance comparison of multiple k-means clustering runs in terms of Sum of Squared Errors (SSE). Once fixed the optimal number of clusters (C = 100), the Davies-Bouldin index (DB) is computed and reported as “DB_C100”. Lower DB values indicate better clustering.

**Supplementary Fig. 2.**
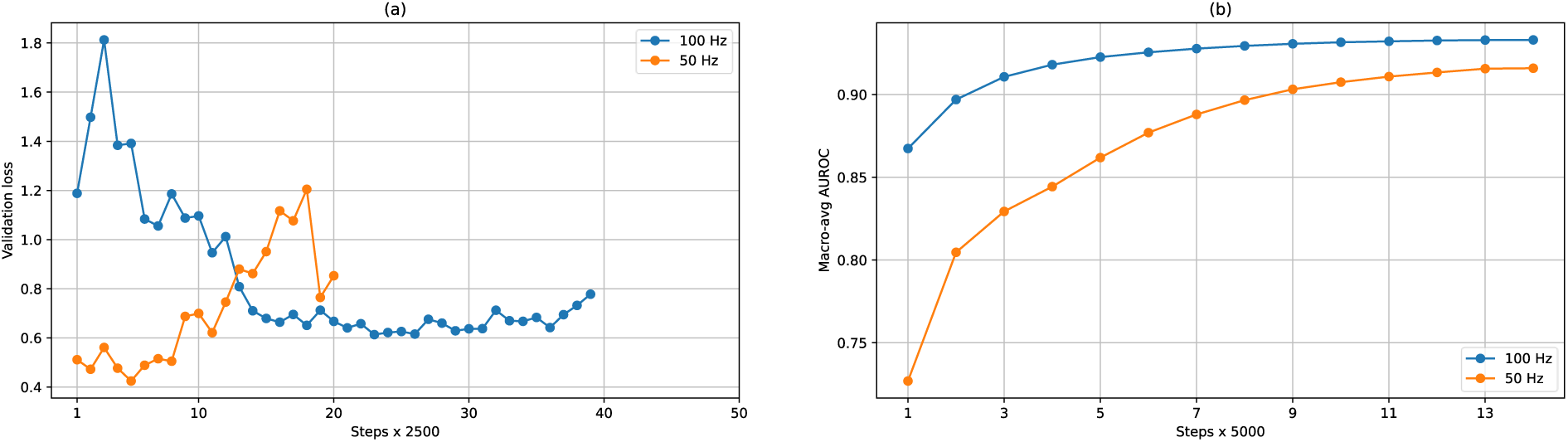
Pre-training and linear evaluation performance with varying sampling rate. **(a)** Validation loss curves obtained when pre-training HuBERT-ECG with ECGs sampled at 50 and 100 Hz. **(b)** Downstream performance obtained when HuBERT-ECG is linearly evaluated on *Ribeiro-dev* after being pre-trained with ECGs sampled at 50 and 100 Hz.

**Supplementary Fig. 3.**
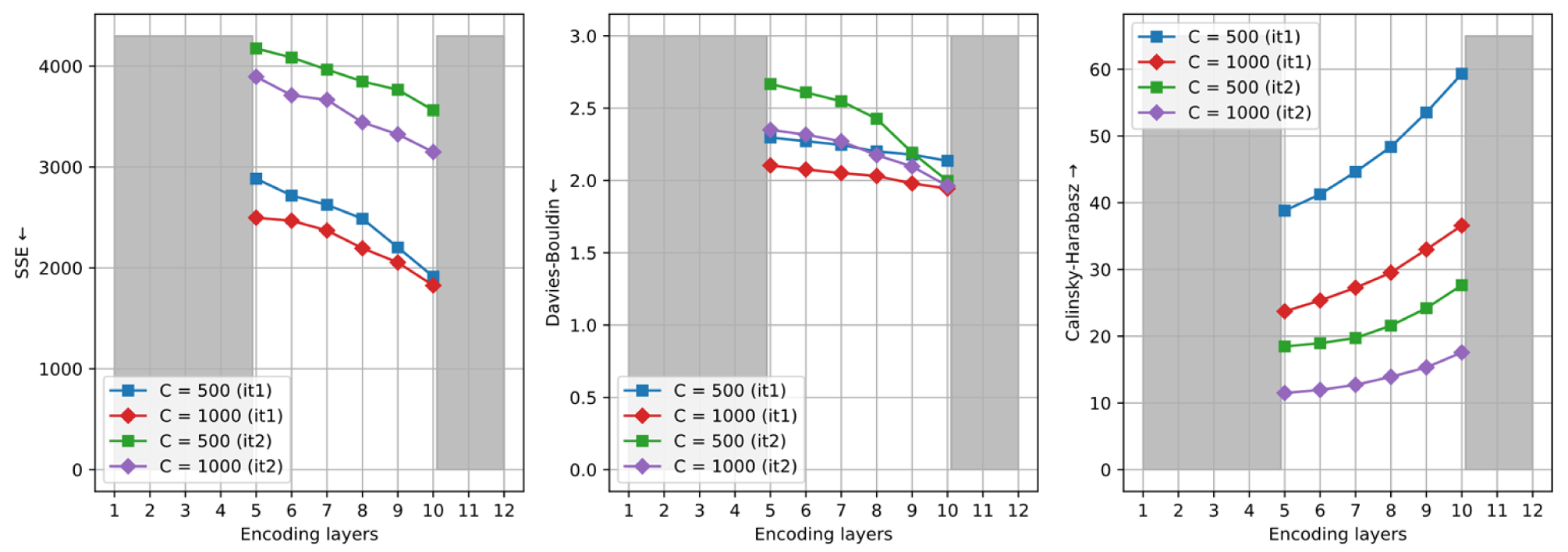
Clustering quality metrics (inertia, Davies-Bouldin index, Calisky-Harabasz index) across HuBERT-ECG BASE encoding layers after the first (it1) and second (it2) pre-training iterations. Symbols ↑and ↓ indicate whether a metric needs to be maximised or minimised, respectively. Gray-shaded regions refer to encoding layers not considered in this analysis.

**Supplementary Fig. 4.**
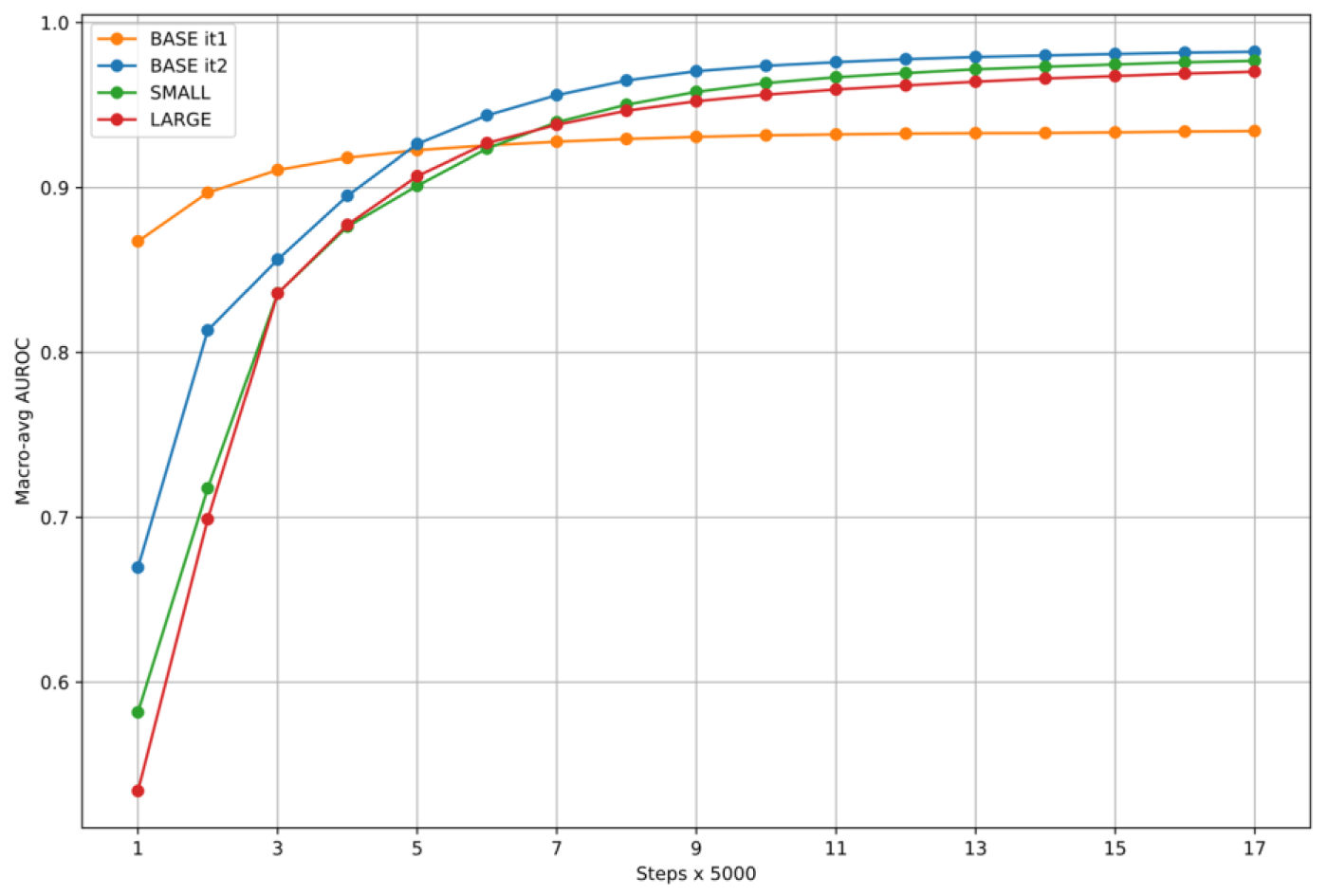
Linear evaluation performance of HuBERT-ECG BASE, after first and second pre-training iteration, HuBERT-ECG SMALL and HuBERT-ECG LARGE on *Ribeiro-dev*.

**Supplementary Fig. 5.**
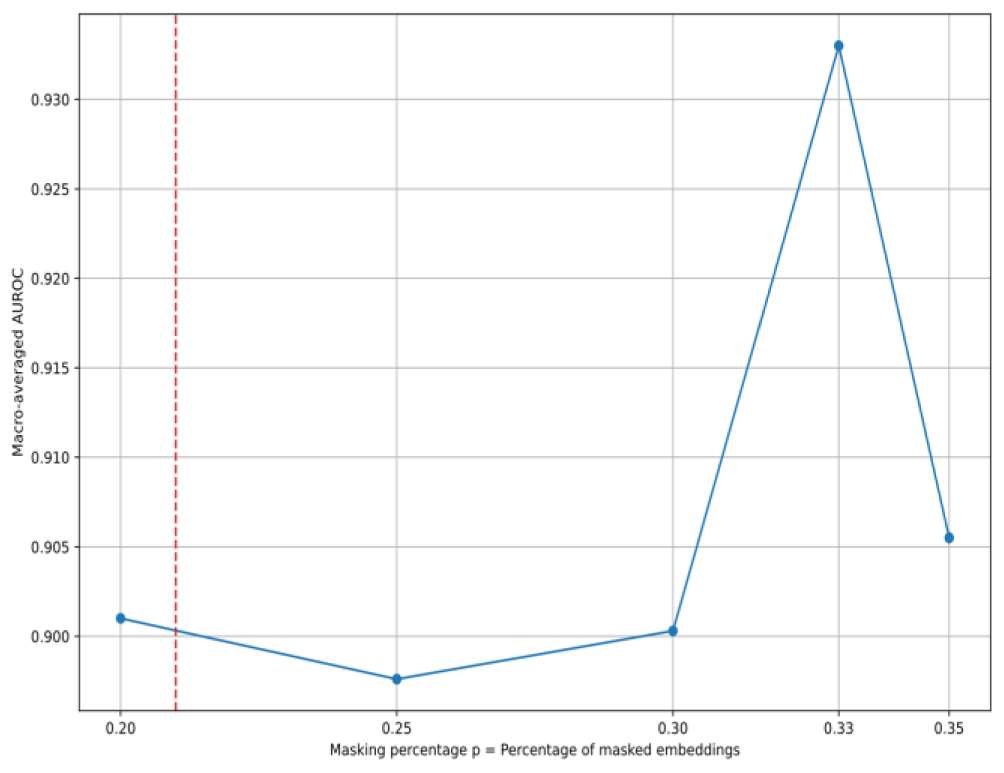
HuBERT-ECG BASE linear evaluation performance after pre-training with different values of the hyper-parameter *p* (i.e. the percentage of ECG embeddings to mask). The red dashed line indicates the percentage of ECG embeddings that would be masked if we followed the masking strategy used to pre-train HuBERT^3^.

**Supplementary Fig. 6.**
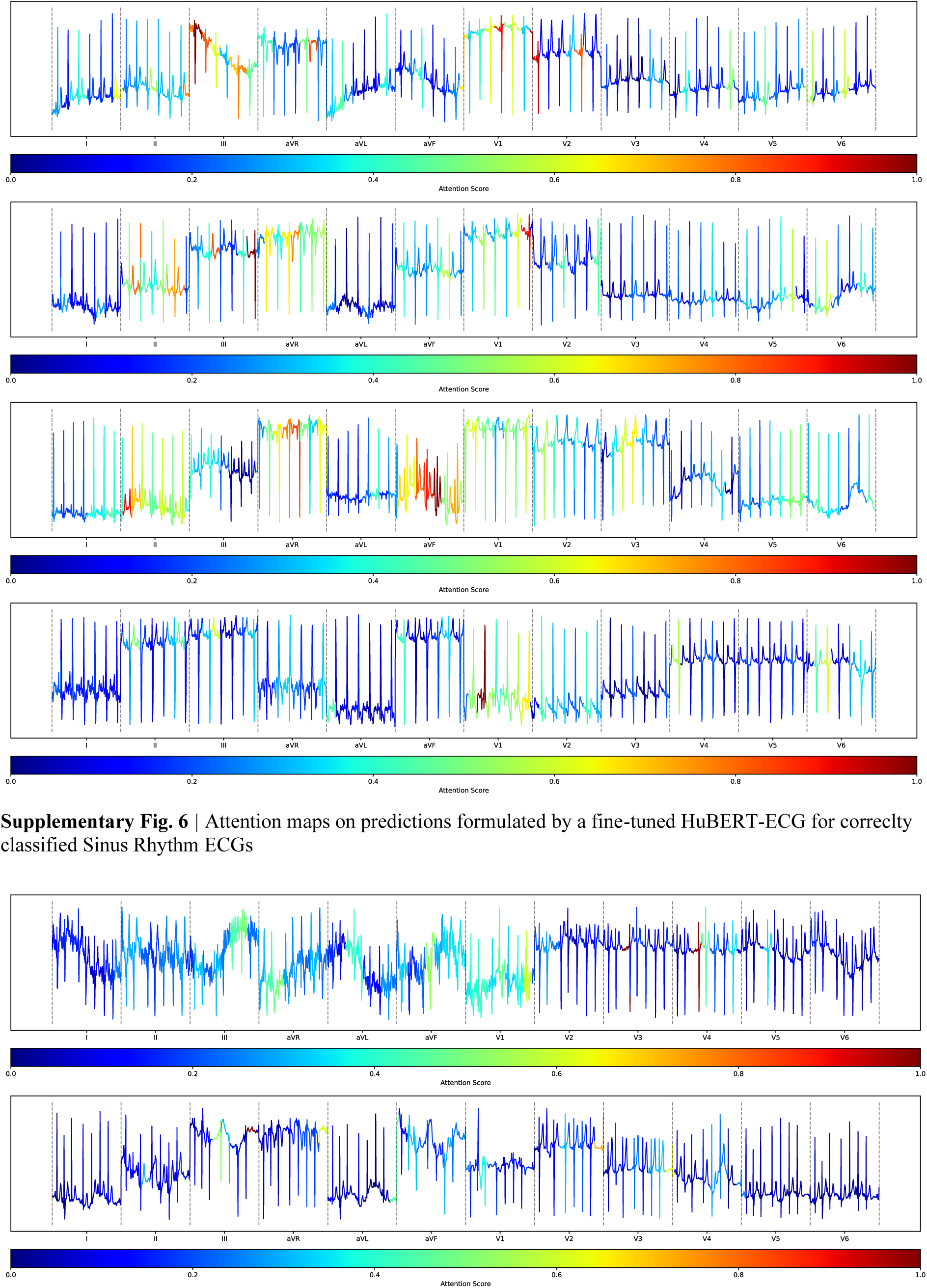
Attention maps on predictions formulated by a fine-tuned HuBERT-ECG for correclty classified Sinus Rhythm ECGs

## Supplementary Tables

**Supplementary Table 1:**
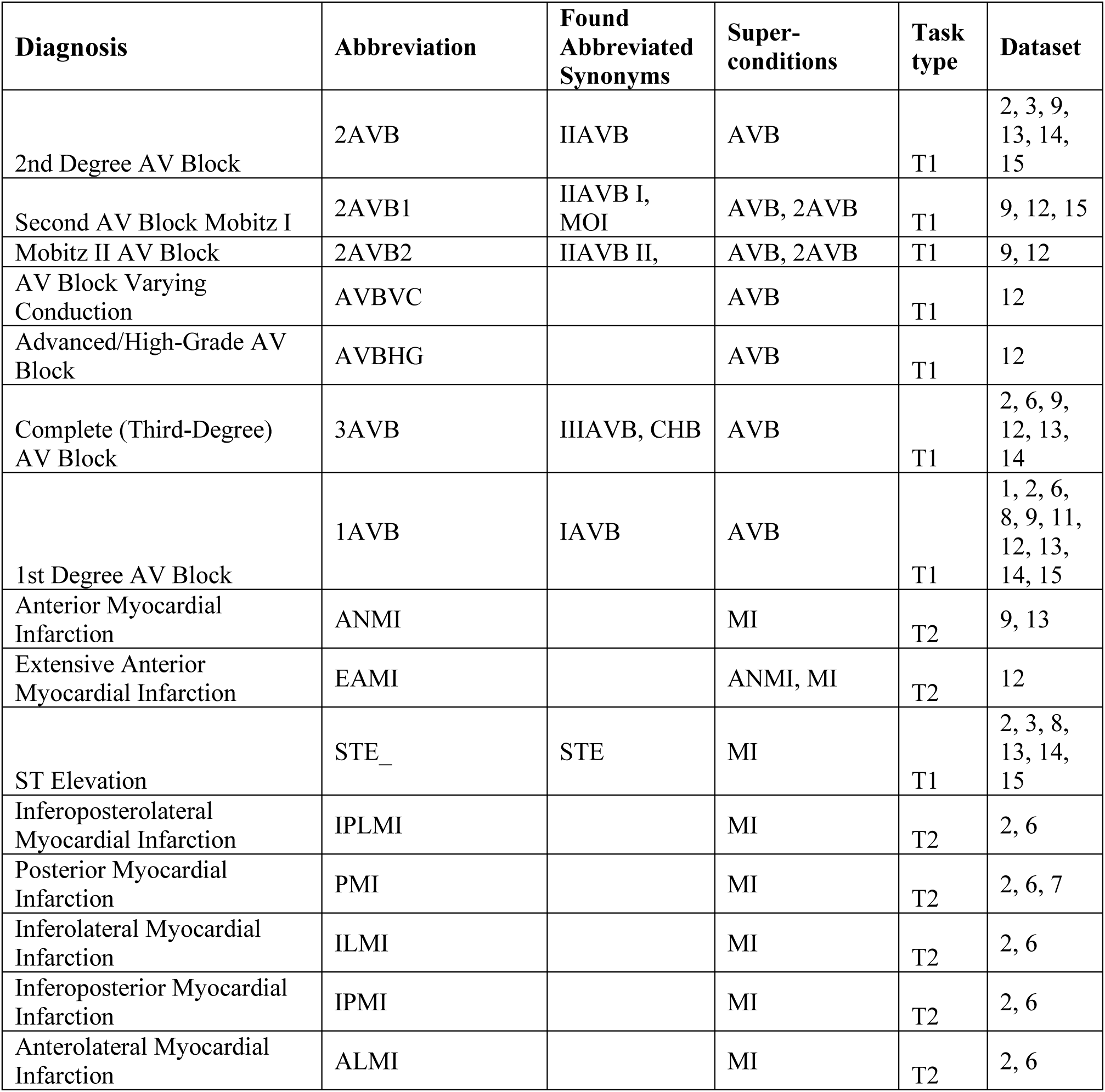
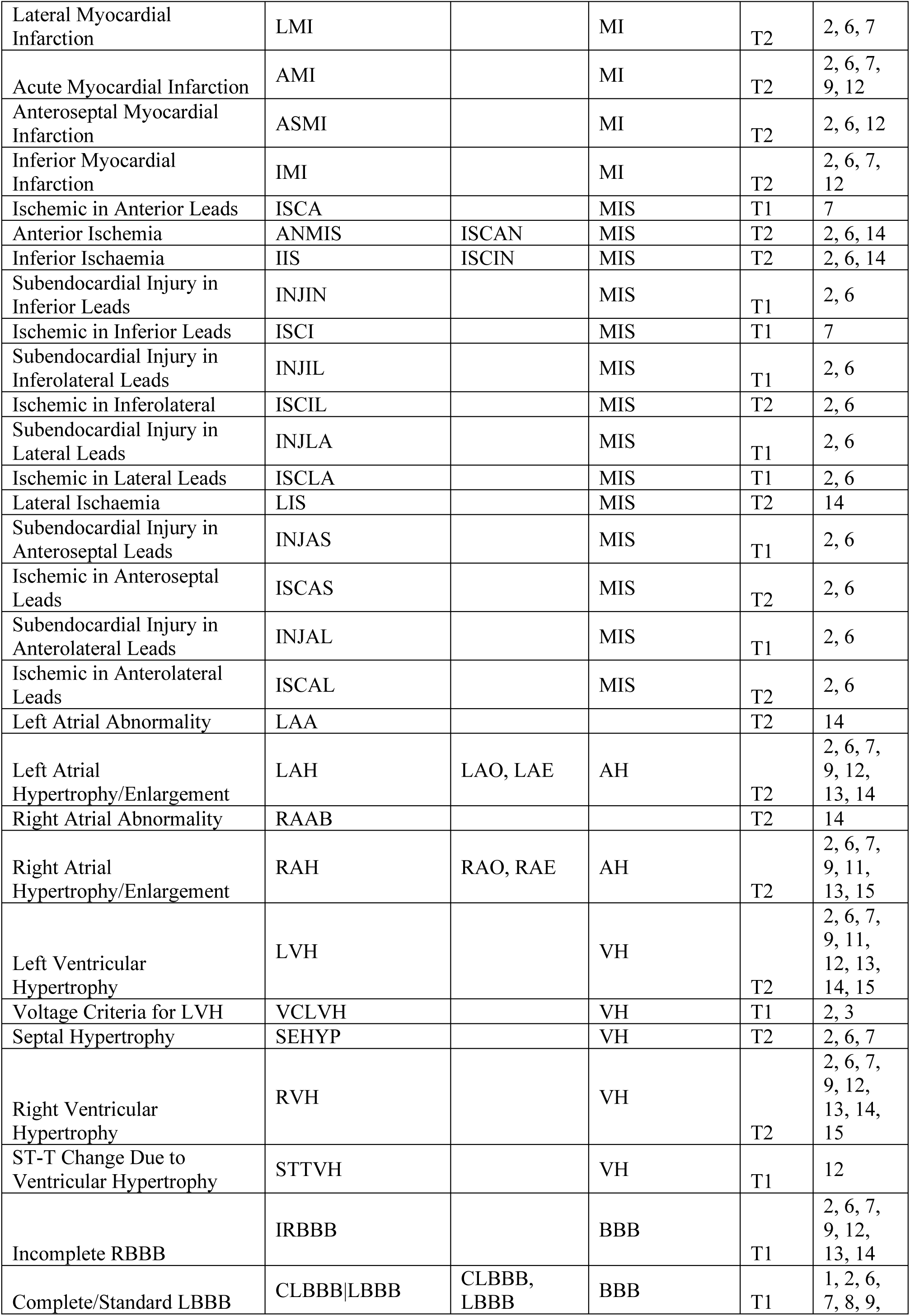
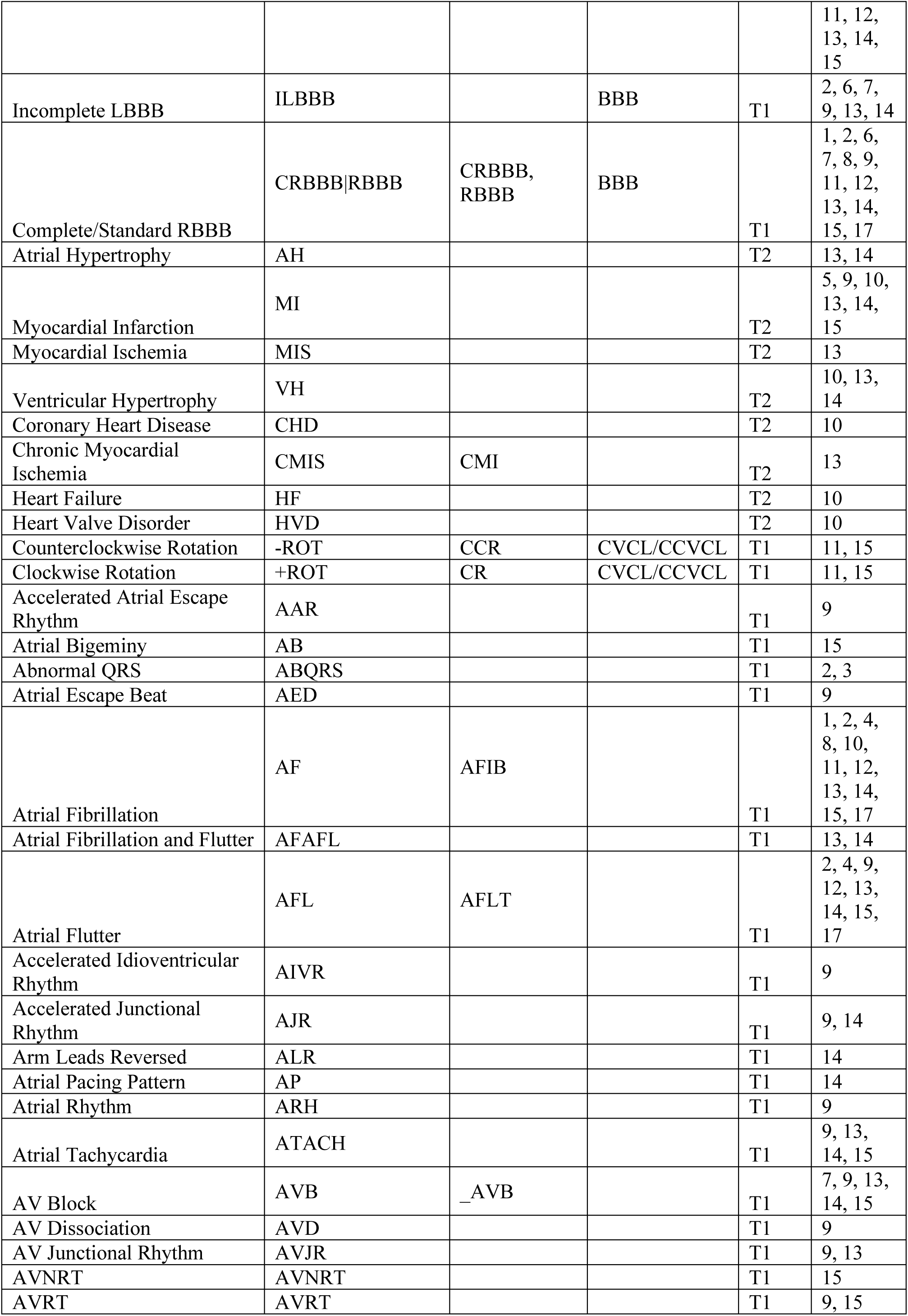
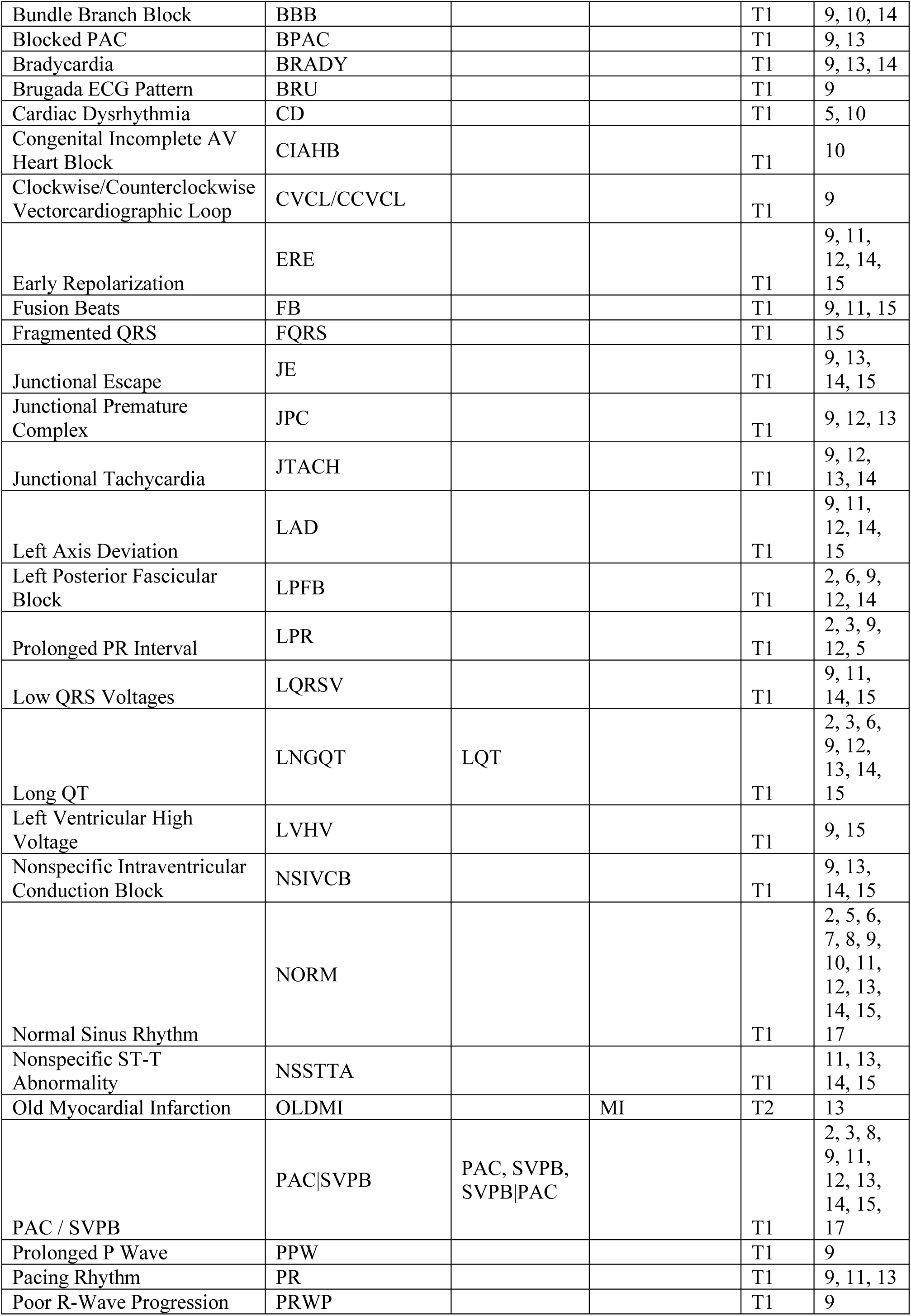
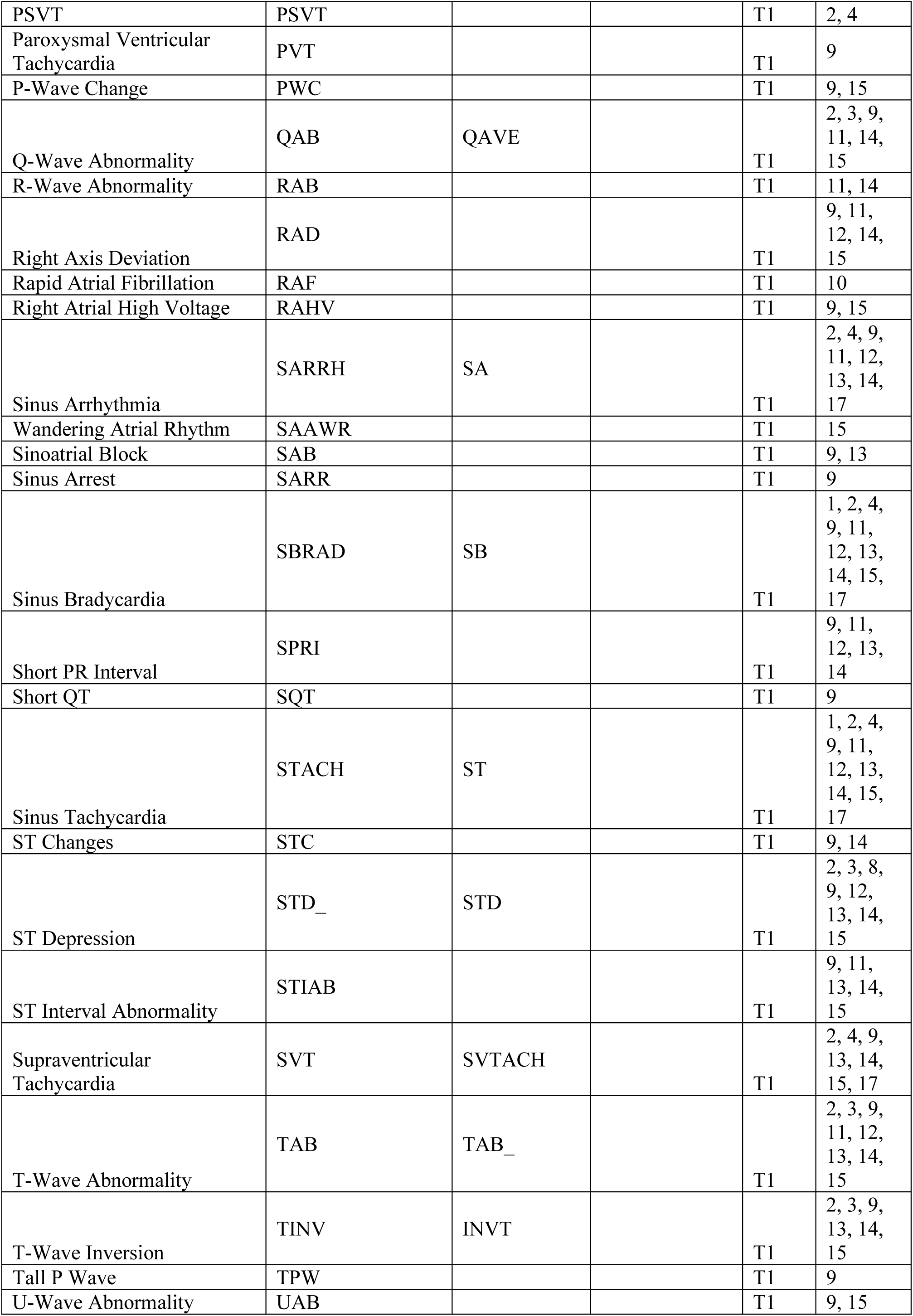
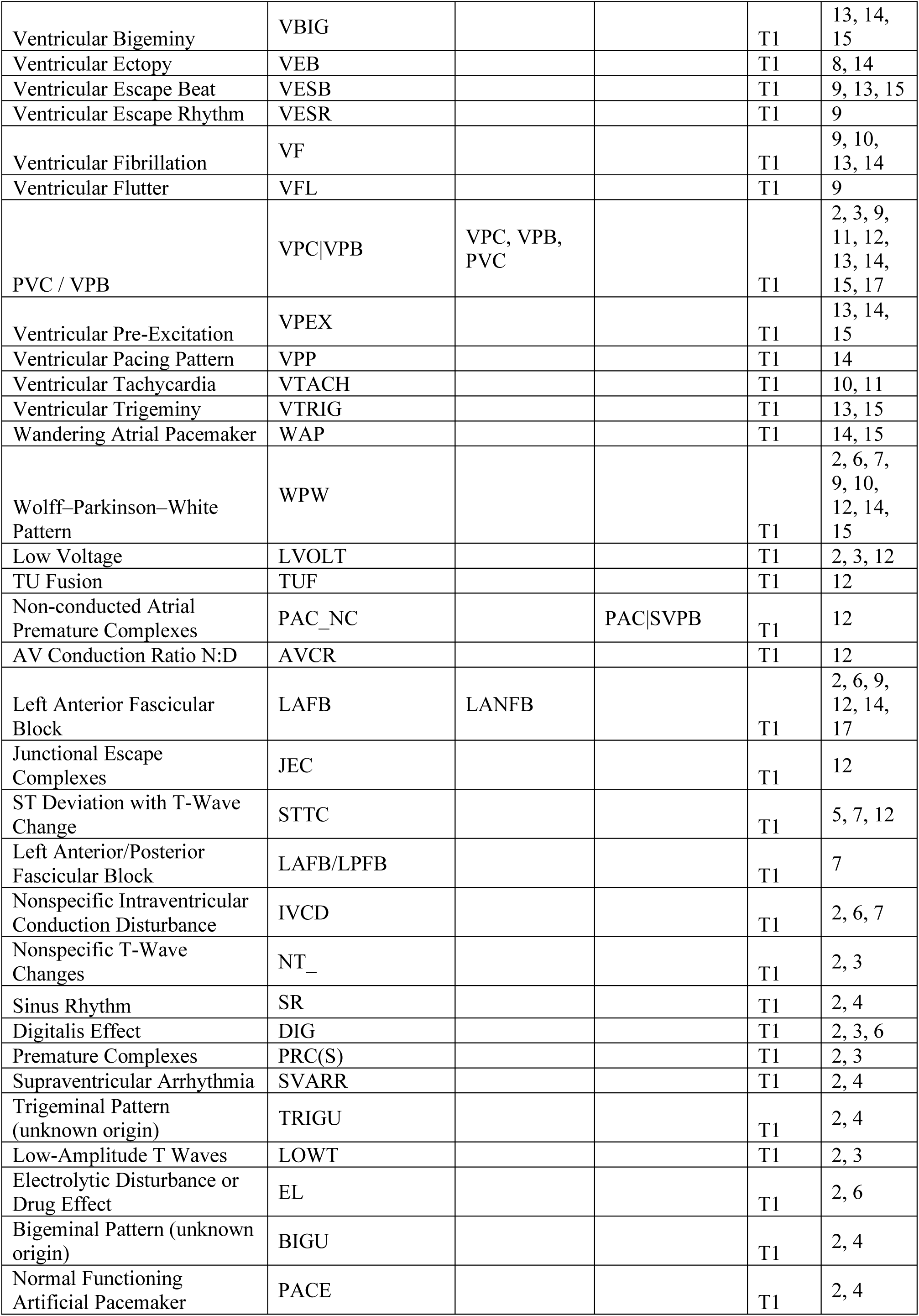
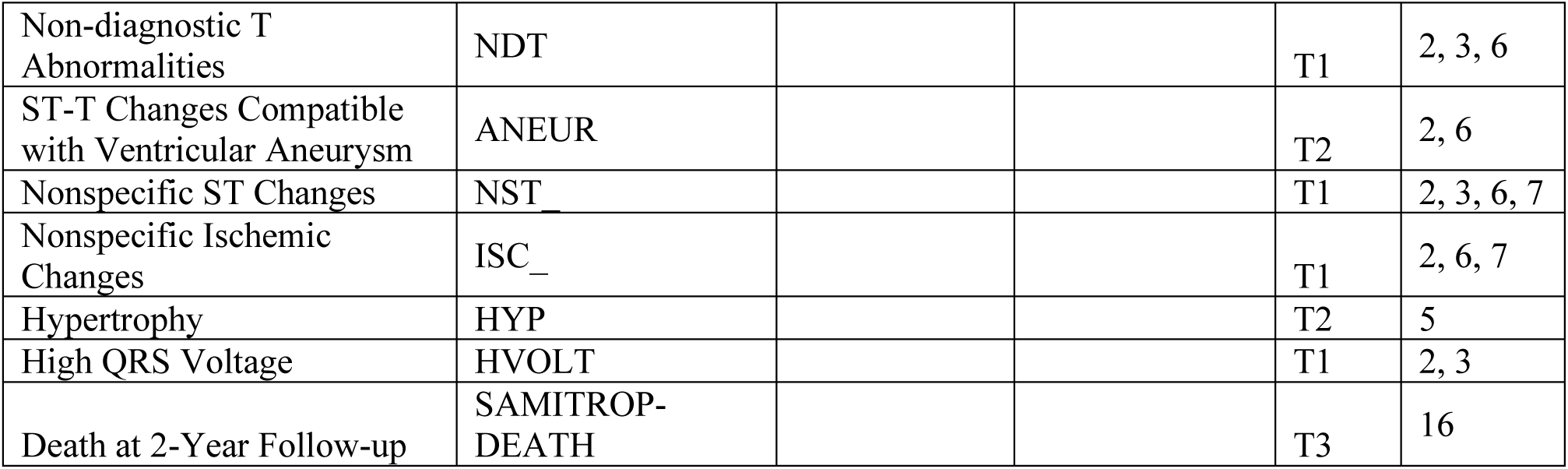
This table lists the harmonised diagnoses and the corresponding abbreviations used throughout this study. For conditions/tasks that appear with different names across various datasets, the table also includes found synonyms. When generating the Cardio-Learning dataset, conditions appearing under various synonyms are standardised to a unique name. The ‘Super-conditions’ column indicates hierarchically broader diagnostic categories. When creating the Cardio-Learning dataset, if broader and more specific conditions come from different constituent datasets, the broader conditions augment the annotation of an ECG that is labelled with the more specific condition. If an ECG is labelled with a more general condition, its annotation is not augmented with a more specific diagnostic category, unless explicitly confirmed in the original annotation. Importantly, these hierarchical augmentations are performed only when the hierarchy involves diagnostic labels coming from different constituent datasets. This means that if a more specific condition and its broader diagnostic category appear in the same dataset, but an ECG from that source is explicitly labelled only the more specific one, the hierarchical mapping is not performed to respect the distinction made by the original annotators. The ‘Dataset’ column lists the datasets in which a certain task is present: 1 stands for Ribeiro, 2-7 stand for PTB-XL (All, Form, Rhythm, Diag. Superclass, Diag., Diag. Subclass), 8 stands for CPSC, 9 stands for Ningbo, 10 stands for PTB,11 stands for Hefei, 12 stands for SPH, 13 stands for CPSC Extra, 14 stands for Georgia, 15 stands for Chapman, 16 stands for SaMi-Trop, 17 stands for “Clinical data”. The ‘Task type’ column reports task type membership based on the diagnostic role of the ECG: T1 (type-1), T2 (type-2), T3 (type-3).

**Supplementary Table 2:** EchoNext’s underlying tasks with corresponding abbreviation and task-type categorization.

| **EchoNext** |  |  |
| --- | --- | --- |
| **Diagnosis** | **Abbreviation** | **Task type** |
| Lower ventricular ejection fraction $\leq 45\%$ | lvwt_gte_13_flag | T2 |
| Interventricular septum or posterior wall thickness $\geq 13\%$ | lvwt_gte_13_flag | T2 |
| Moderate or severe aortic stenosis | aortic_stenosis_moderate_or_greater_flag | T2 |
| Moderate or severe aortic regurgitation | aortic_regurgitation_moderate_or_greater_flag | T2 |
| Moderate or severe mitral regurgitation | mitral_regurgitation_moderate_or_greater_flag | T2 |
| Moderate or severe tricuspid regurgitation | tricuspid_regurgitation_moderate_or_greater_flag | T2 |
| Moderate or severe pulmonary regurgitation | pulmonary_regurgitation_moderate_or_greater_flag | T2 |
| Moderate or severe right ventricular systolic dysfunction. | rv_systolic_dysfunction_moderate_or_greater_flag | T2 |
| Presence of moderate or large pericardial effusion | pericardial_effusion_moderate_large_flag | T2 |
| pulmonary artery systolic pressure $\geq 45\text{mmHG}$ | pasp_gte_45_flag | T2 |
| tricuspid regurgitation maximum velocity $\geq 32\text{ cm/s}$ | tr_max_gte_32_flag | T2 |
| Composite binary label indicating presence of a | shd_moderate_or_greater_flag | T2 |
| structural heart disease |  |  |

**Supplementary Table 3:** MITHSDB’s classes and task-type categorization. Being this dataset used for multi-class classification, there is only one underlying task of type 2, since the gold-standard for diagnosis is not the ECG (e.g. echocardiography, phonocardiography, doppler imaging, or hemodynamics).

| MITHSDB |  |  |
| --- | --- | --- |
| Diagnosis | Abbreviation | Task type |
| Normal heart sound | NORM HS | T2 |
| Murmur in systole | MVP |  |
| Mitral Regurgitation | MR |  |
| Mitral Stenosis | MS |  |
| Aortic stenosis | AS |  |

**Supplementary Table 4:** Linear evaluation performance of pre-trained HuBERT-ECG BASE models when using pseudo-labels generated by MFCC- and time-frequency based k-means models.

| Label generator | Macro-averaged AUROC |
| --- | --- |
| MFCC-based k-means – C = 100 | 0.933 |
| Time-frequency based k-means – C = 100 | 0.913 |

**Supplementary Table 5:**
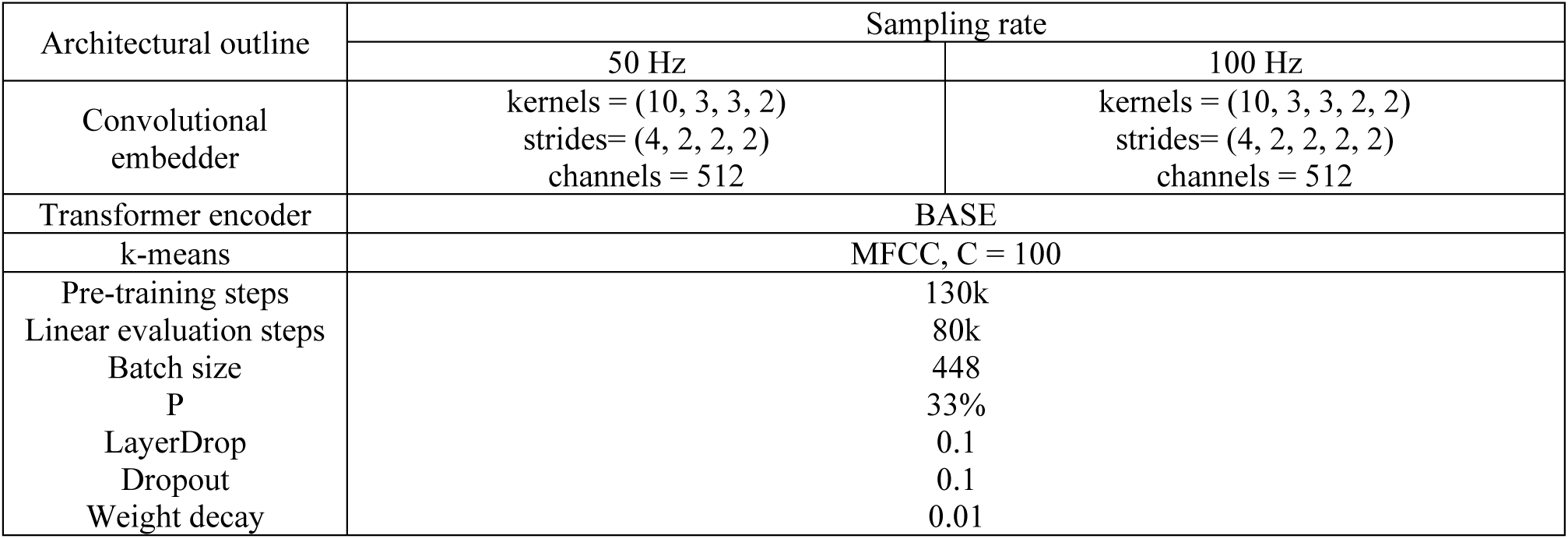
Architecture design and running configuration when experimenting with ECGs sampled at 50 and 100 Hz.

**Supplementary Table 6:** Linear evaluation performance of HuBERT-ECG on *Ribeiro-dev* after being pre-trained at 50 and 100 Hz. Linear evaluation steps necessary to let validation AUROC plateau are also reported.

| ECG sampling rate | macro-averaged AUROC | Linear evaluation steps at plateau |
| --- | --- | --- |
| 50 Hz | 0.9176 | 65k |
| 100 Hz | 0.9330 |  |

**Supplementary Table 7:**
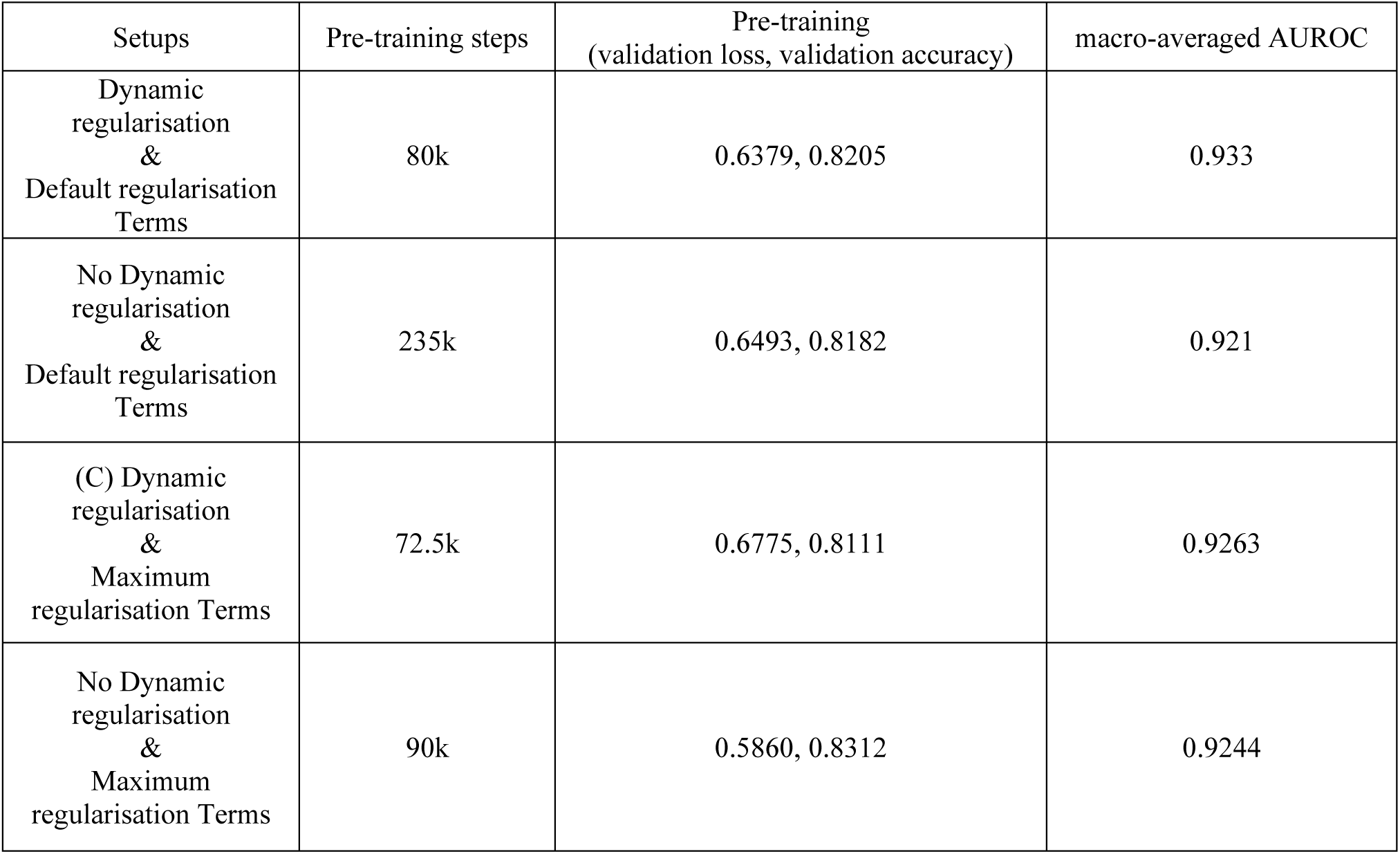
Effects of the use of dynamic regularisation when pre-training HuBERT-ECG in terms of pre-training steps and upstream performance under the form of validation loss and validation accuracy. For each setup, linear evaluation performance on *Ribeiro-dev* is also reported.

**Supplementary Table 8:** Linear evaluation performance of HuBERT-ECG BASE after being pre-trained in a multi-task learning framework in which tasks are represented by multiple k-means models composing an ensemble of label generators. The corresponding number of pre-training steps is reported for every task/cluster ensemble.

| K-means ensemble | Macro-averaged AUROC | Pre-training steps |
| --- | --- | --- |
| MFCC-based k-means, C = 100 | 0.933 | 80k |
| MFCC-based k-means, C = 100, 200 | 0.930 | 75k |
| MFCC-based k-means, C = 100, 200, 300 | 0.943 | 135k |

**Supplementary Table 9:** Comparison of all HuBERT-ECG configurations with existing models from the literature on Ribeiro test set according to multiple metrics. Sensitivity and specificity are obtained setting the threshold to 0.5, as done by Ribeiro et al., for a fair comparison. Performance uncertainty is reported using 95% confidence intervals computed via empirical bootstrapping (N=1000). Label abbreviations and corresponding diagnosis can be found in Supplementary Table 1.

| Condition | Models | Sensitivity | Specificity | AUROC | AUPRC |
| --- | --- | --- | --- | --- | --- |
| AF | Ribeiro et al. | 0.769 | 1.000 | 0.885 | 0.773 |
|  | ECGFounder | 1.000 | 0.996 | 0.999 | N.A. |
|  | HuBERT-ECG SMALL | 1.0000<br>(CI:0.9999–1.0000) | 0.9754<br>(CI:0.9656–0.9853) | 1.0000<br>(CI:0.9986–1.0000) | 0.9774<br>(CI:0.9278–1.0000) |
|  | HuBERT-ECG BASE | 1.0000<br>(CI:0.999–1.000) | 1.0000<br>(CI:0.9999–1.0000) | **1.0000**<br>(CI:0.9999–1.0000) | **1.0000**<br>(CI:0.9999–1.0000) |
|  | HuBERT-ECG LARGE | 1.0000<br>(CI:0.9999–1.000) | 0.9748<br>(CI:0.9644–0.9853) | 0.9976<br>(CI:0.9954–0.9999) | 0.9240<br>(CI:0.8535–0.9945) |
| IAVB | Ribeiro et al. | 0.929 | 0.995 | 0.962 | 0.807 |
|  | ECGFounder | 0.864 | 0.957 | 0.949 | N.A. |
|  | HuBERT-ECG SMALL | 0.9290<br>(CI:0.8580–1.0000) | 0.9531<br>(CI:0.9387–0.9675) | 0.9890<br>(CI:0.9817–0.9963) | 0.8309<br>(CI:0.7366–0.9252) |
|  | HuBERT-ECG BASE | 0.7500<br>(CI:0.6071–0.8929) | 0.9981<br>(CI:0.9962–1.0000) | **0.9977**<br>(CI:0.9961–0.9994) | **0.9560**<br>(CI:0.9264–0.9856) |
|  | HuBERT-ECG LARGE | 1.0000<br>(CI:0.9990–1.0000) | 0.9485<br>(CI:0.9337–0.9637) | 0.9960<br>(CI:0.9935–0.9985) | 0.8750<br>(CI:0.7929–0.9571) |
| CLBBB\|LBBB | Ribeiro et al. | 1.000 | 1.000 | 1.000 | 1.000 |
|  | ECGFounder | 1.000 | 0.996 | 0.998 | N.A. |
|  | HuBERT-ECG SMALL | 1.0000<br>(CI:0.9999–1.0000) | 0.9940<br>(CI:0.9893–0.9987) | 1.0000<br>(CI:0.9999–1.0000) | 0.9950<br>(CI:0.9837–1.0000) |
|  | HuBERT-ECG BASE | 0.9000<br>(CI:0.8333–0.9667) | 1.0000<br>(CI:0.9999–1.0000) | **1.0000**<br>(CI:0.9999–1.0000) | **1.0000**<br>(CI:0.9999–1.0000) |
|  | HuBERT-ECG LARGE | 0.9330<br>(CI:0.8993–0.9667) | 0.9950<br>(CI:0.9913–0.9987) | 0.9983<br>(CI:0.9971–0.9995) | 0.9760<br>(CI:0.9609–0.9911) |
| CRBBB\|RBBB | Ribeiro et al. | 1.000 | 0.995 | 0.997 | 0.895 |
|  | ECGFounder | 0.971 | 0.971 | 0.989 | N.A. |
|  | HuBERT-ECG SMALL | 1.0000<br>(CI:0.9999–1.000) | 0.9850<br>(CI:0.9776–0.9924) | **0.9987**<br>(CI:0.9976–0.9998) | 0.9800<br>(CI:0.9616–0.9984) |
|  | HuBERT-ECG BASE | 0.9400<br>(CI:0.8529–1.0000) | 0.9943<br>(CI:0.9899–0.9987) | 0.9986<br>(CI:0.9974–0.9998) | **0.9740**<br>(CI:0.9513–0.9967) |
|  | HuBERT-ECG LARGE | 0.9710<br>(CI:0.9420–1.0000) | 0.9874<br>(CI:0.9798–0.9950) | 0.9977<br>(CI:0.9957–0.9998) | 0.9690<br>(CI:0.9424–0.9956) |
| SBRAD | Ribeiro et al. | 0.938 | 0.996 | 0.967 | 0.782 |
|  | ECGFounder | 1.000 | 0.955 | 0.967 | N.A. |
|  | HuBERT-ECG SMALL | 1.0000<br>(CI:0.9999–1.0000) | 0.9808<br>(CI:0.9716–0.9901) | 0.9973<br>(CI:0.9948–0.9998) | 0.8801<br>(CI:0.7719–0.9883) |
|  | HuBERT-ECG BASE | 1.0000<br>(CI:0.9999–1.0000) | 0.9919<br>(CI:0.9864–0.9975) | **0.9980**<br>(CI:0.9954–1.0000) | 0.8860<br>(CI:0.7504–1.0000) |
|  | HuBERT-ECG LARGE | 1.0000<br>(CI:0.9999–1.0000) | 0.9796<br>(CI:0.9704–0.9889) | 0.9980<br>(CI:0.9962–0.9998) | **0.8950**<br>(CI:0.8014–0.9886) |
| STACH | Ribeiro et al. | 0.937 | 0.997 | 0.985 | 0.923 |
|  | ECGFounder | 0.974 | 0.970 | 0.989 | N.A. |
|  | HuBERT-ECG SMALL | 0.9730<br>(CI:0.9729–0.9731) | 0.9816<br>(CI:0.9722–0.9911) | 0.9969<br>(CI:0.9946–0.9992) | 0.9350<br>(CI:0.8807–0.9893) |
|  | HuBERT-ECG BASE | 0.9730<br>(CI:0.9729–0.9731) | 0.9950<br>(CI:0.9913–0.9987) | **0.9990**<br>(CI:0.9983–0.9997) | **0.9640**<br>(CI:0.9342–0.9938) |
|  | HuBERT-ECG LARGE | 0.9730<br>(CI:0.9728–0.9732) | 0.9829<br>(CI:0.9734–0.9924) | 0.9957<br>(CI:0.9939–0.9975) | 0.9480<br>(CI:0.9149–0.9811) |
| MACRO-AVERAGED | Ribeiro et al. | 0.935 | 0.997 | 0.966 | 0.863 |
|  | ECGFounder | 0.9682 | 0.9742 | 0.9818 | N.A. |
|  | HuBERT-ECG SMALL | 0.9806<br>(CI:0.9657–0.9955) | 0.9784<br>(CI:0.9740–0.9828) | 0.9974<br>(CI:0.9964–0.9984) | 0.9330<br>(CI:0.9045–0.9615) |
|  | HuBERT-ECG BASE | 0.9255<br>(CI:0.8943–0.9567) | 0.9969<br>(CI:0.9954–0.9983) | **0.9989**<br>(CI:0.9983–0.9995) | **0.9630**<br>(CI:0.9403–0.9857) |
|  | HuBERT-ECG LARGE | 0.9770<br>(CI:0.9641–0.9899) | 0.9782<br>(CI:0.9740–0.9825) | 0.9978<br>(CI:0.9973–0.9983) | 0.9310<br>(CI:0.9013–0.9607) |

**Supplementary Table 10:** Fine-tuned HuBERT-ECG performance on PTB-XL datasets against models from the literature. Performance uncertainty is reported using 95% confidence intervals obtained through empirical bootstrapping using 1000 bootstrap iterations. “N.A.” stands for “Not Available”. Results preceded by ‘∼’ symbol are estimated by looking at paper graphs because of the lack of unequivocal numerical values.

| Models | PTB-XL<br>All | PTB-XL<br>Form | PTB-XL<br>Rhythm | PTB-XL<br>Diag. | PTB-XL<br>Diag.<br>Subclass | PTB-XL<br>Diag.<br>Superclass |
| --- | --- | --- | --- | --- | --- | --- |
|  | Macro-averaged AUROC |  |  |  |  |  |
| HuBERT-ECG<br>SMALL | 0.905±0.006 | 0.844±0.012 | 0.946±0.013 | 0.931±0.005 | 0.917±0.005 | 0.909±0.007 |
| HuBERT-ECG<br>BASE | 0.911±0.002 | 0.855±0.012 | 0.959±0.010 | 0.941±0.005 | 0.918±0.006 | 0.911±0.008 |
| HuBERT-ECG<br>LARGE | 0.900±0.003 | 0.834±0.018 | 0.942±0.016 | 0.925±0.002 | 0.923±0.006 | 0.908±0.007 |
| Hu et al.<sup>15</sup> | 0.947 | ~ 0.895 | ~ 0.980 | ~ 0.950 | ~ 0.940 | ~ 0.938 |
| Liu et al.<sup>16</sup> | N.A. | N.A. | N.A. | N.A. | N.A. | 0.912 |
| Liu et al.<sup>17</sup> | N.A. | N.A. | N.A. | N.A. | N.A. | 0.877 |
| Mehari & Strodthoff<sup>18</sup> | 0.942 | N.A. | N.A. | N.A. | N.A. | N.A. |
| Strodthoff et al.<sup>19</sup> | 0.925 | 0.896 | 0.957 | 0.937 | 0.929 | 0.928 |
| Na et al.<sup>20</sup> | N.A. | N.A. | N.A. | N.A. | N.A. | 0.933 |
| Zhang et al.<sup>21</sup> | 0.889 | N.A. | N.A. | N.A. | N.A. | N.A. |
| Zhou et al.<sup>22</sup> | N.A. | N.A. | N.A. | N.A. | N.A. | 0.932 |

## Footnotes

a https://tianchi.aliyun.com/competition/entrance/231754/introduction

b https://tianchi.aliyun.com/competition/entrance/231754/introduction

c PTB-XL generates 6 different datasets that differ from each other in the presented conditions.

d We express cross-entropy loss as a simple sum of log probabilities, instead of the traditional sum over *p*(·)*log*(*p*(·)), because, in masked cross-entropy, *p*(·) terms are implicitly equal to 1, given the one-hot encoding of labels.

e https://wandb.ai/site

f https://huggingface.co/Edoardo-BS

## Notes

### Competing Interest Statement

The authors have declared no competing interest.

### Summary of Updates

New results and findings on acute-care prediction, long and short-term prognostic modelling, ECG-based echocardiographic finding prediction, patient characteristics regression, and transferability to single-lead ECG settings typical of portable devices. New results are summarised are reported in the text and summarised in new Tables and Figures (e.g., new Fig. 4).

